# Striatal Dopaminergic Asymmetry as a marker of Brain-First and Body-First Subtypes in de novo Parkinson’s Disease

**DOI:** 10.1101/2022.12.23.22283910

**Authors:** Jeffrey M Boertien, Muhammad Nazmuddin, Justyna Kłos, Anne C Slomp, Sygrid van der Zee, Ronald JH Borra, Teus van Laar

**Author notes:** **Corresponding author:** Jeffrey Boertien, University Medical Center Groningen, Department of Neurology (AB51), P.O. Box 30.001, 9700RB Groningen, The Netherlands. Jeffrey M Boertien and Muhammad Nazmuddin should be considered joint first authors. **Trial registration:** NCT04180865 (DUPARC).

## Abstract

Recently, the α-Synuclein Origin and Connectome (SOC) model of Parkinson’s disease (PD) has been proposed, which predicts a more malignant clinical subtype and symmetrical neurodegeneration in body-first compared to brain-first PD.

Here, motor symptoms (MDS-UPDRS III), non-motor symptoms (NMSQ) and T1 MRI data of an incident *de novo* PD cohort, were compared between PD subjects with levels of putaminal dopaminergic asymmetry in the lowest tertile (PD-sym, n=41) and highest tertile (PD-asym, n=41), as measured by FDOPA-PET.

PD-sym was associated with a higher burden of motor symptoms and non-motor symptoms with a probable neurological substrate caudally from the substantia nigra. Though overall brain volume was lower in PD-sym, no differences in the volumes and asymmetricity of specific brain regions could be found between PD-sym and PD-asym after adjusting for multiple testing.

The more malignant clinical picture suggests an overrepresentation of body-first PD subjects in PD-sym according to the SOC-model. Also, lower overall brain volumes were found in PD-sym. However, structural MRI data might not be sufficient to assess regional differential degeneration between PD-sym and PD-asym in *de novo* PD. Additional imaging modalities and longitudinal follow-up could be required to support or reject the SOC-model.

## Introduction

Parkinson’s disease (PD) is the second most common neurodegenerative disorder, characterized by a large variety of motor and non-motor symptoms.^1^ Due to the clinical heterogeneity of PD, various attempts have been made to create PD subtypes.^2^ Most prominently, clinical PD subtypes have been defined according to the motor symptomatology into tremor-dominant (TD) and postural instability and gait disorder (PIGD) subtypes, or into clusters of non-motor symptomatology.^3,4^ Though previous classifications might explain some variability in disease progression and clinical presentation, they do not adequately address possible etiological differences.

Concerning the etiology of PD, the most accepted paradigm describes PD as a prion-like disorder in which alpha-synuclein (aSyn) aggregation leads to self-propagation and neuro-anatomical spreading of Lewy body pathology.^5^ According to Braak, PD pathology might begin in the periphery, entering the brain via either the dorsal motor nucleus of the vagal nerve or the olfactory bulb.^6,7^ In line with Braak’s hypothesis, PD is often preceded by a prodromal phase characterized by non-motor symptoms such as REM sleep behavior disorder (RBD), constipation and orthostatic hypotension.^8^ In particular RBD has a high predictive value with a pheno-conversion rate to an alpha-synucleïnopathy of more than 70% after 12 years.^9^ In addition, Lewy body pathology has been found in the gut wall of prodromal PD subjects, years before diagnosis.^10^ Nonetheless, Braaks’ hypothesis does not account for the large clinical variability observed in PD. In particular, the existence of PD subjects who have not experienced prodromal symptomatology and the different cognitive profiles of PD subjects.

Recently, the α-Synuclein (aSyn) Origin and Connectome (SOC) Model has been proposed to explain the clinical heterogeneity of PD within the framework of prion-like propagation.^11^ The main foundation for the SOC-model is the finding that RBD-positive PD subjects, as well as RBD-positive probable prodromal PD subjects, show more denervation along the vagal nerve compared to RBD-negative PD subjects.^12^ A dichotomy based on the site of origin of aSyn pathology is proposed, suggesting a probable body-first (pPD-body) and brain-first (pPD-brain) subtype. Subsequently, clinical variability can be explained by different spreading patterns along neuroanatomical connections. In particular, asymmetry of neurodegeneration would be more pronounced in the pPD-brain subtype as intra-hemisphere connections greatly outnumber inter-hemisphere connections.^13^ For a gastrointestinal origin, as most likely pPD-body site-of-onset, rodent studies show cross-linked neuroanatomical connections within the enteric nervous system and along the vagal nerve.^14,15^ This would lead to a more symmetrical burden of aSyn pathology in both hemispheres at the time of diagnosis and subsequently a more malignant pPD-body subtype. In concordance, RBD-positive PD was found to have a more symmetric dopaminergic deficit compared to RBD-negative PD at the time of diagnosis, before a floor effect is reached when the striatal dopaminergic projections become depleted.^16,17^ Asymmetricity of the dopaminergic deficit at the time of diagnosis might therefore be a suitable proxy to enrich populations with pPD-brain and pPD-body subjects according to the SOC-model.

Several concrete hypotheses have been formulated to assess the SOC-model of PD. These include (1) more pronounced neurodegeneration due to more symmetrical pathology in pPD-body; (2) more pronounced non-motor symptomatology in pPD-body, in particular of non-motor symptoms with a neurologic substrate caudal from the substantia nigra (SN) (e.g., autonomic dysfunction); (3) faster cognitive decline in pPD-body; (4) asymmetric neurodegeneration in pPD-brain, mainly limited to the hemisphere with the largest dopaminergic deficit; and (5) more neurodegeneration of limbic structures in the most affected hemisphere of pPD-brain, as limbic structures, in particular the amygdala and entorhinal cortex, have been proposed as possible sites-of-origin of brain-first Lewy body disorders.^11,18,19^

In the current study the above hypotheses were assessed using clinical and structural (T1) MRI data of treatment-naïve *de novo* PD subjects. PD subjects were dichotomized according to the asymmetry of the putaminal dopaminergic deficit as measured by 3,4-dihydroxy-6-18F-fluoro-1-phenylalaninie (FDOPA) PET scan imaging, with the lowest and highest tertile representing symmetrical and asymmetrical PD, relatively enriched with pPD-body and pPD-brain, respectively, according to the SOC-model.

## Results

### Participant selection

128 DUPARC participants with complete FDOPA-PET and T1-MRI data from the Dutch Parkinson Cohort of de novo PD subjects (DUPARC) were included.^20^ Participants whose absolute values of the striatal asymmetricity index (SAI) of the FDOPA-PET striatal-to-occipital ratios (SOR) of the putamen were in the highest and lowest tertiles, were classified as asymmetrical PD (PD-asym) and symmetrical PD (PD-sym) respectively.^21^

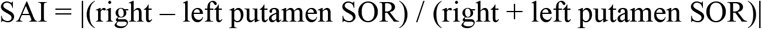

One PD-asym participant was removed after quality control of MRI data. To balance both groups, one PD-sym participant was removed, resulting in 41 participants per group. Putaminal SORs were similar between PD-sym and PD-asym on the most affected side, but were higher in PD-asym on the least affected side (*p=*3.06e-10, Figure 1). SAIs of putamen and caudate showed a strong correlation, but the range was twice as large in the putamen, supporting its use to differentiate between PD-sym and PD-asym (Supplementary Figure 1).

**Figure 1.**
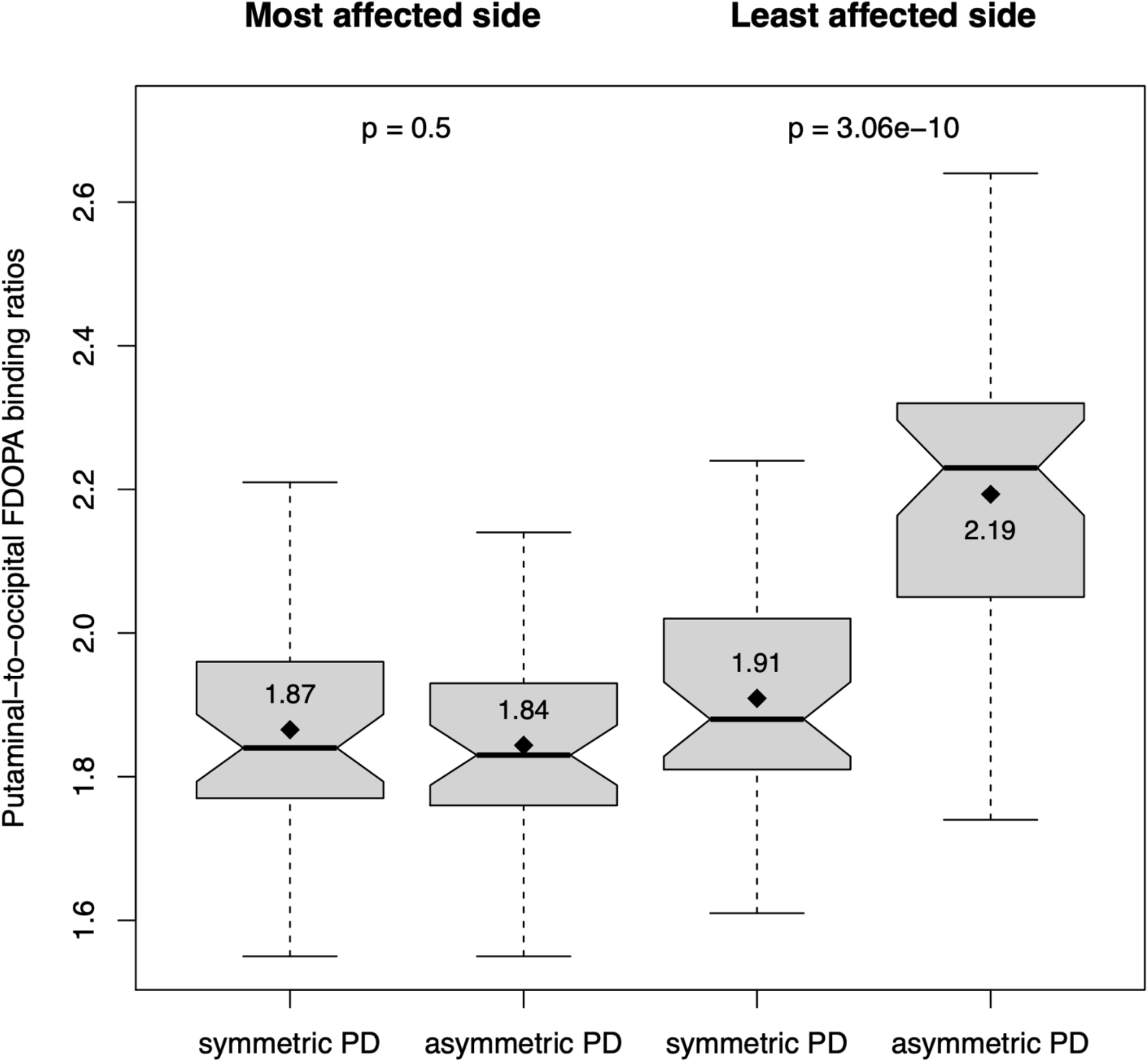
Putaminal striatal-to-occipital FDOPA binding ratios in symmetric and asymmetric Parkinson’s disease (PD). Boxplots represent the median with the first and third quartile. Whiskers represent values up to 1.5 times the interquartile range. Diamonds represent the mean value, which are written in the boxplot. The left two boxplots are on the most affected side, whereas the right two boxplots are on the least affected sides. A statistically significant difference in putaminal striatal-to-occipital binding ratios was found at the least affected side (*p=*3.06E-10) whereas no difference was found at the most affected side (*p=*0.5).

### Clinical characteristics

PD-sym and PD-asym represent two clinically distinct groups. PD-sym is characterized by a higher age and a higher motor symptom burden as measured by the Movement Disorders Society Unified Parkinson’s Disease Rating Scale (MDS-UPDRS) part III and the Hoehn and Yahr stage.^22^ Moreover, participants were classified as tremor dominant (TD), postural instability and gait disorder (PIGD) or indeterminate subtypes.^3^ PD-sym was also characterized by a larger proportion of the PIGD subtype (Table 1). Overall non-motor symptom burden, as measured by the Movement Disorders Society Non-Motor Symptom Questionnaire (NMSQ) total scores, did not show a statistically significant difference. An additional analysis was performed only including NMSQ items with a probable predominant neurological substrate inferior to the substantia nigra (SN), as judged by a movement disorders specialist (see Methods). NMSQ items classified as anatomically originating caudal to the SN were more prevalent in PD-sym. No difference was found between the non-motor symptoms hyposmia and RBD, using Sniffin’ sticks and the RBD screening questionnaire (RBDSQ), respectively. In particular, the scores for question 6 did not differ between groups, although it has a higher sensitivity for RBD in *de novo* PD than the RBDSQ total score.^23^ Cognitive screening by means of the Montreal Cognitive Assessment (MoCA) showed worse performance in PD-sym (*p=*0.032), but this difference was not statistically significant after correction for multiple testing. No differences were found in the male to female ratio and level of education. Also, no differences were found in self-reported motor symptom duration, suggesting no differences in diagnostic delay between the two groups. An overview of the clinical characteristics of PD-sym and PD-asym is provided in table 1.

**Table 1.** Clinical Characteristics.

|  | **Symmetric PD**<br>**(n=41)** | **Asymmetric PD**<br>**(n=41)** | **p-value** | **p-FDR** |
| --- | --- | --- | --- | --- |
| **Age**<br>Mean (SD) | 68.5 (7.83) | 63.0 (8.16) | **0.0023** | **0.015** |
| **Sex** %Female | 26.8% | 29.3% | 1 | 1 |
| **Education<sup>a</sup>**<br>Median [IQR] | 5 [4-6] | 5 [4-6] | 0.41 | 0.56 |
| **MDS-UPDRS III** Mean<br>(SD) | 36.0 (12.64) | 28.3 (10.52) | **0.0038** | **0.016** |
| **Hoehn & Yahr**<br>Median [IQR] | 2 [2-2] | 2[1-2] | **0.0015** | **0.015** |
| **Motor subtype**<br>PIGD:IND:TD% | 66:10:24 | 34:15:51 | **0.015** | **0.039** |
| **Motor symptom**<br>**duration (months)<sup>b</sup>**<br>Median [IQR] | 24 [12-24] | 24 [12-30] | 0.49 | 0.58 |
| **NMSQ-Total**<br>Median [IQR] | 6 [4-9] | 5 [2-8] | 0.056 | 0.10 |
| **Below-SN NMSQ**<br>**Score<sup>c</sup>** Median [IQR] | 5 [3-6] | 3 [1-5] | **0.011** | **0.036** |
| **RBDSQ-Total**<br>Median [IQR] | 4 [2-6] | 3 [1-6] | 0.40 | 0.56 |
| **RBDSQ Question 6<sup>d</sup>**<br>Median [IQR] | 1 [0-1] | 0 [0-2] | 0.53 | 0.58 |
| **MoCA score**<br>Mean (SD) | 24.1 (3.04) | 25.5 (2.81) | **0.032** | 0.068 |
| **Sniffin'stick score**<br>Mean (SD) | 5.1 (2.56) | 5.5 (2.13) | 0.43 | 0.56 |

Abbreviations: HC, healthy control; PD, Parkinson's disease; TD, tremor dominant; PIGD, postural instability and gait difficulty; IND, indeterminate; MDS-UPDRS-III, Movement Disorder Society- Unified Parkinson's Disease Rating Scale–Motor Subscale; MoCA, Montreal Cognitive Assessment; NMSQ, Nonmotor Symptoms Questionnaire; SN, substantia nigra; RBDSQ, REM sleep behavior disorders screening questionnaire.
<sup>a</sup>Educational level according to the Dutch Verhage Scale (Verhage, 1964). <sup>b</sup>n=35 for symmetric PD and n=37 for asymmetric PD. <sup>c</sup>Total scores of NMSQ items assessing non-motor-symptoms associated with structures caudal to substantia nigra (item 1, 3-9, 19, 20, 24-26, 28, 29). <sup>d</sup>Subscore question 6 of the RBDSQ, consisting of the sum total of 6A, 6B, 6C, 6D.

### Brain atrophy

T1-weighted MRI scans were processed using the cNeuro software version 1.11.0 (Combinostics Oy, Tampere, Finland, www.cneuro.com). Percentile scores of volumes of predetermined brain regions were provided, based on an internal reference dataset of 1923 control subjects (18-94 years old, 57% female), normalized for age, sex and head size. cNeuro was used because it is an automated clinical-grade brain MRI quantification solution, allowing for direct translation of potential findings into clinical practice. The scans were inspected with automated cNeuro signal quality control. Additionally, a visual quality control of the derived structural cNeuro segmentation was performed. Hemispheres were labeled as most or least affected relative to the dopaminergic deficit after calculation of the percentile scores, thereby accounting for pre-existent dexterity.

PD-sym showed lower percentile scores of total brain volume compared to PD-asym (*p=*0.029), which was associated with a relative loss of cerebral white matter volume (*p=*0.0034) rather than cerebral gray matter volume (*p=*0.39) (Figure 2, Supplementary Table 1). Of the assessed individual brain regions, the cerebellar vermal lobules VIII-X and the supramarginal gyrus showed statistically significant higher percentile scores in PD-sym compared to PD-asym before correction for multiple testing. In contrast, the percentile scores of the pallidum and hippocampus were lower in PD-sym compared to PD-asym before correction for multiple testing. However, after false discovery rate (FDR) correction for multiple testing, none of the assessed individual brain regions showed a statistically significant difference between the two groups.

**Figure 2.**
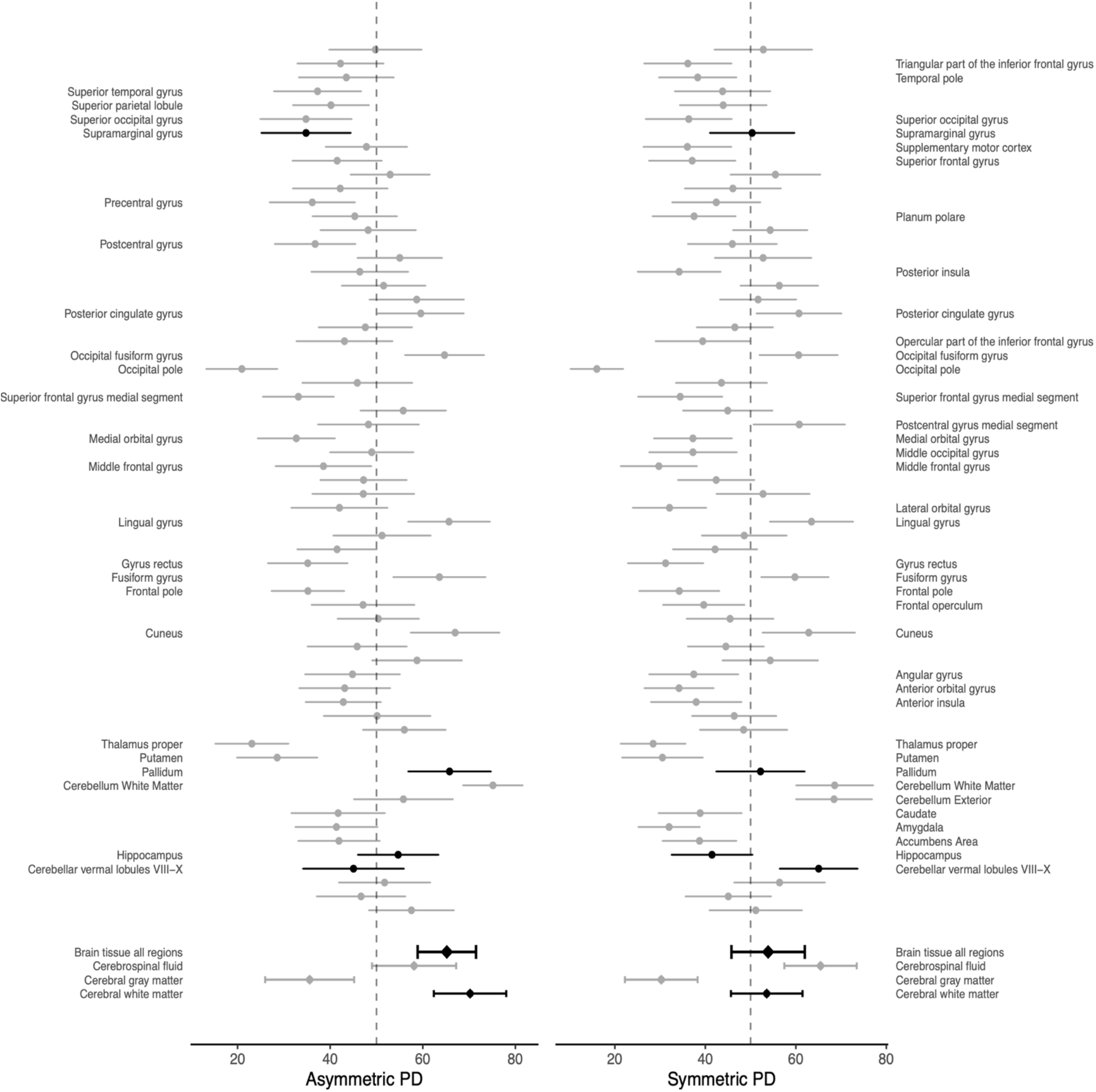
Mean percentile scores of brain region sizes of symmetric and asymmetric Parkinson’s disease (PD). Depicted are the mean percentile scores of brain region size, as calculated by cNeuro, with their 95% confidence interval (CI). Larger values represent larger brain region size as compared to the control population, normalized for age, sex and head size. Highlighted are brain regions with a statistically significant difference in percentile scores between both groups before correction for multiple testing. None of the brain regions differed with statistical significance after correction for multiple testing (FDR). In addition, brain areas of which the CI does not include the 50^th^ percentile are annotated. Total brain tissue of all regions, cerebrospinal fluid, cerebral gray matter and cerebral white matter were included as overall measures of brain volume. Asymmetric PD is characterized by a higher total brain tissue volume (*p=*0.029), mainly driven by higher white matter percentile scores (*p=*0.0034).

In addition, voxel-wise analyses were applied to compare gray matter volume between groups. Prior to applying these analyses, MRIs of PD subjects with dominant left hemispheric dopaminergic deficit were flipped using FSL’s fslswapdim (https://fsl.fmrib.ox.ac.uk/fsl/fslwiki/). Voxel-based morphometry (VBM) analysis of gray matter structures were performed based on previously described standardized pipelines using the Statistical Parametric Mapping software version 12 (SPM12).^24,25^ Results were adjusted in separate analyses for both age and total intracranial volume (TIV) or only for TIV.

The VBM analysis also showed no differences in gray matter density between PD-sym and PD-asym when adjusting for TIV and age at FDR p<0.05. When only adjusted for TIV, gray matter density was decreased in PD-sym in 28 structures in the most and least affected hemispheres at FDR p<0.05 (Figure 3, Supplementary Table 2). Relative to PD-asym, more and larger areas of gray matter density loss were found in the least affected hemisphere, compared to the most affected hemisphere. Areas belonging to the caudate, amygdala, insula, inferior temporal gyrus and inferior frontal gyrus pars triangularis were identified as smaller in PD-sym in both hemispheres. No areas with decreased gray matter density in PD-asym relative to PD-sym were identified.

**Figure 3.**
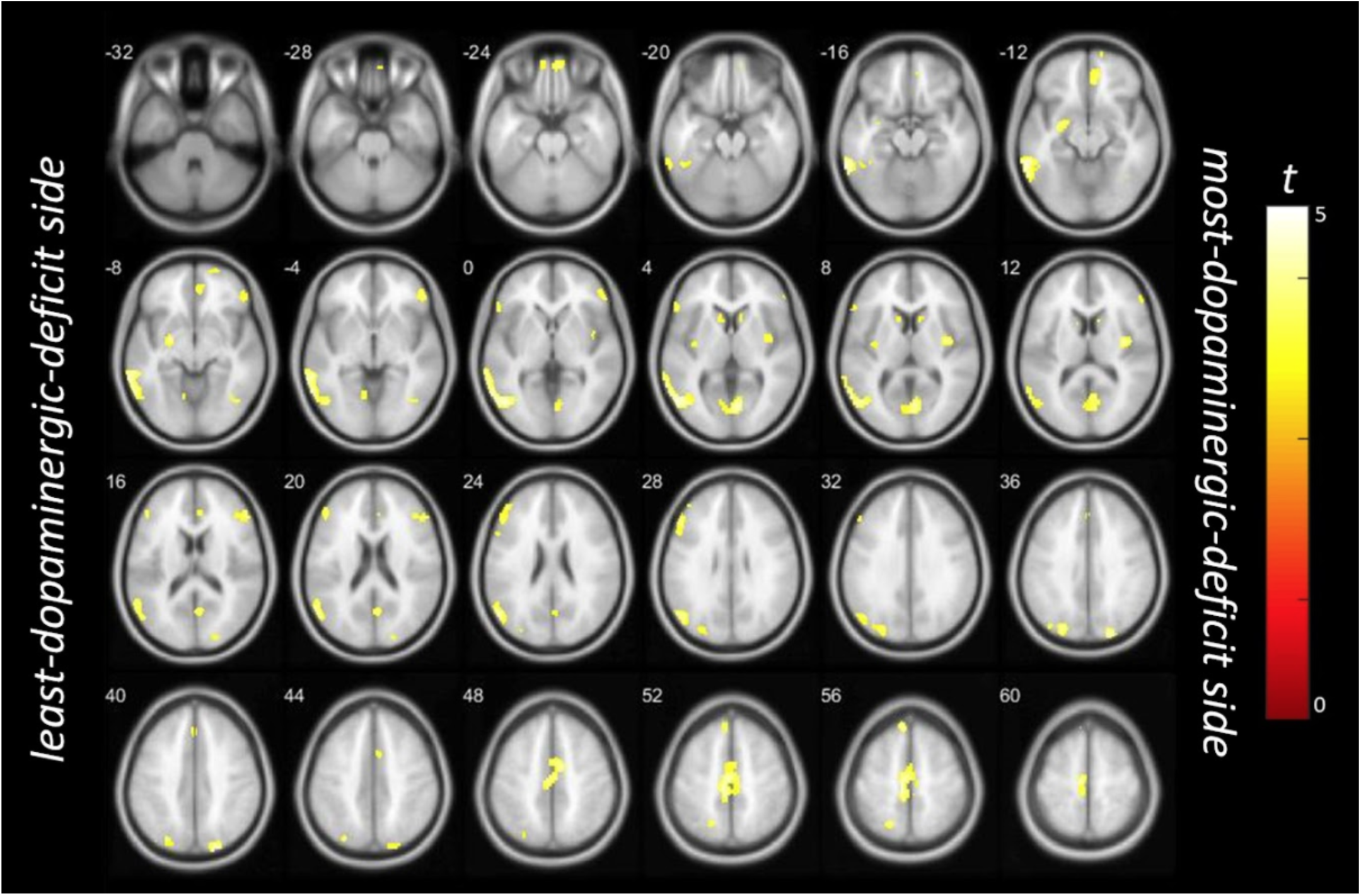
Clusters of significant gray matter atrophy (FDR p < 0.05, k = 50), revealed by voxel-based morphometry analysis adjusted for intracranial volume, in symmetric PD compared to the asymmetric PD. MRI data were pooled as if the right brain hemisphere is the side with the most putaminal dopaminergic depletion as measured by F-DOPA PET.

### Brain region asymmetry

The SOC-model predicts more asymmetric brain atrophy in PD-asym, mainly limited to the hemisphere with the largest dopaminergic deficit. Therefore, brain region asymmetricity indices (BAI) were compared between both groups:

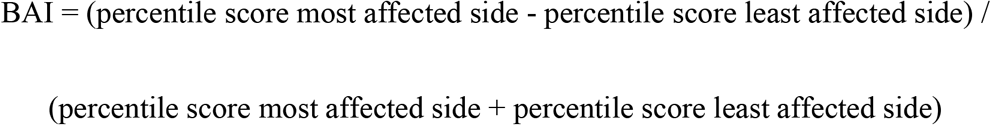

A statistically significant difference was found for the BAI of the transverse temporal gyrus, middle temporal gyrus and posterior cingulate gyrus between PD-sym and PD-asym before correction for multiple testing (Figure 4, Supplementary Table 3). All with a relative negative BAI in PD-asym, indicating lower percentile scores on the most affected hemisphere relative to the dopaminergic deficit. However, none of the identified brain areas differed between both groups after FDR correction. Also, no statistically significant difference was found in the BAI of overall cerebral gray and white matter. PD-asym was characterized by five brain regions with a mean negative BAI that does not include 0 in the 95% CI: the transverse temporal gyrus, superior frontal gyrus, posterior cingulate gyrus, entorhinal area and the thalamus (Figure 4). The precuneus was the only brain region in PD-asym with a positive BAI. PD-sym was characterized by a positive BAI for the transverse temporal gyrus, the middle frontal gyrus and the cerebellum exterior and a negative BAI for the anterior cingulate gyrus.

**Figure 4.**
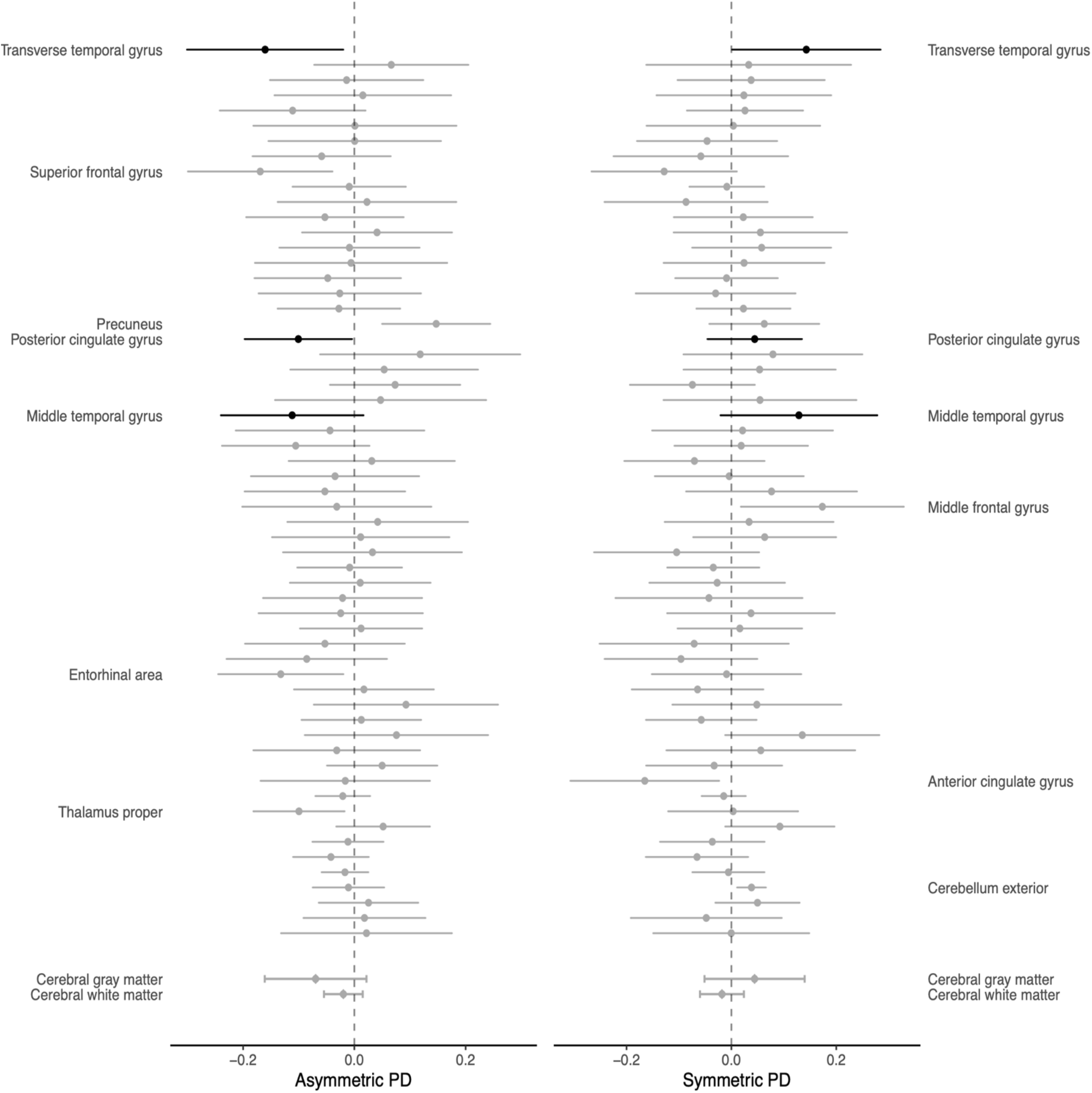
Asymmetry indices of indicated brain regions in asymmetric and symmetric Parkinson’s disease (PD). Depicted are the mean brain region asymmetry indices (BAIs) and associated 95% confidence intervals (CI). Negative values represent smaller brain region size in the most affected hemisphere relative to the least affected hemisphere according to the dopaminergic deficit. Highlighted are brain regions with a statistically significant difference between both groups before correction for multiple testing. In addition, brain regions of which the CI does not include 0 are annotated. Though more brain regions had a negative BAI in asymmetric PD, none showed a statistically significant difference between both groups after correction for multiple testing (FDR). Also, no statistically significant difference was found between the BAI of cerebral gray and white matter, reported as overall measures of brain asymmetry.

In addition, a voxel-wise asymmetry analysis was performed within two regions-of-interest, the amygdala and the entorhinal cortex, both suggested as possible sites-of-origin in pPD-brain. According to the SOC-model, more pronounced atrophy in the hemisphere with the largest dopaminergic deficit would be expected in PD-asym. However, no statistically significant difference in mean asymmetry of the amygdala and entorhinal cortex was found between PD-sym and PD-asym. Also, no differences were found in the volumes of both areas between the groups, except for a larger mean amygdala volume in the least affected hemisphere of PD-asym (Supplementary Figure 2).

### Guilt-by-association analysis of brain regions asymmetry in PD-asym

BAIs of brain regions in PD-asym were correlated amongst each other, using Pearson correlations, in an exploratory guilt-by-association analysis to identify clusters of brain regions with similar BAI deviations. Euclidean distances of the correlation coefficients were used for hierarchical clustering according to Ward’s method. Pearson correlations of clusters of brain regions identified as highly correlated in PD-asym were also performed in PD-sym to assess whether the found correlations were specific for PD-asym.

Clustering of brain regions with a strong correlation between their BAIs in PD-asym identified a cluster with relative strong correlations when cutting the dendrogram in three clusters. The cluster contained an overrepresentation of (limbic) structures in the temporal lobe: the amygdala, hippocampus, planum temporale, entorhinal area, superior temporal gyrus, inferior temporal gyrus and the temporal pole. However, several structures that were highly correlated in PD-asym were also strongly correlated in PD-sym (Figure 5, Supplementary Table 4).

**Figure 5.**
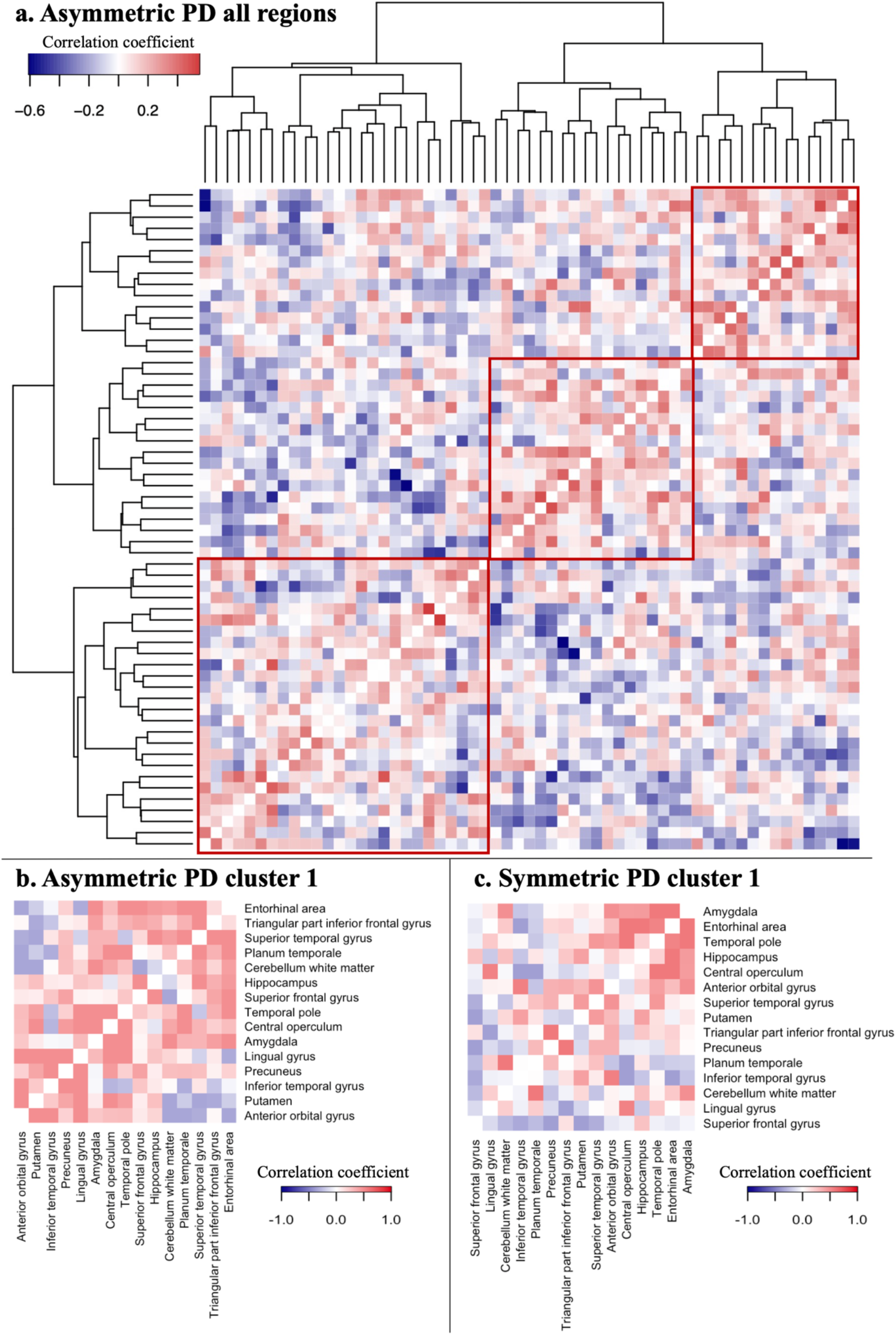
Correlation of brain regions asymmetry indices (BAIs) in asymmetric Parkinson’s disease (PD). (a) Clustering of Pearson correlation coefficients using Ward’s method based on Euclidean distances, reveals a cluster with relative strong positive correlations with an overrepresentation of (limbic) structures in the temporal lobe when cutting the dendrogram in three (highlighted in the red square in the top right corner). No strong overrepresentation of connected brain regions was found in the other two clusters (Supplementary Table 4). (b) Correlation coefficients of the identified cluster with annotated brain regions. (c) Correlation of the identified brain regions in PD-sym also shows strong correlations among several (limbic) structures in the temporal lobe, in particular the amygdala, entorhinal area, temporal pole and hippocampus.

## Discussion

The current study assesses several hypotheses posed by the SOC-model, in relation to the asymmetry of the pathology and associated symptomatology in PD. The assessments were performed in treatment-naïve *de novo* PD subjects not related to the cohort in which the brain-first versus body-first dichotomy was first described.^12^ In concordance with the SOC-model, PD-sym represents a more malignant subtype with more motor symptoms and more non-motor symptomatology of NMSQ-items with a probable predominant neurological substrate below the SN. Also, PD-sym was associated with lower percentile scores of overall brain volume than PD-asym. However, no individual brain region was identified as more atrophic or asymmetric between both groups after FDR-correction. Moreover, global cognitive performance and overall brain asymmetry did not differ between groups.

### Clinical comparison

Despite the absence of a prevalence estimate and asymmetry cut-off, non-motor correlates of pPD-body were found in PD-sym with more non-motor symptoms with a probable predominant neurological substrate below the SN. These mainly concerned autonomic symptoms such as constipation, drooling and orthostatic hypotension. Although hyposmia has been associated with reduced cardiac innervation, suggesting a relation with pPD-body, there was no difference in olfactory function between PD-sym and PD-asym in the current analysis.^26^ The SOC-model predicts faster cognitive decline in pPD-body.^27^ The current study readily indicates a possible differential cognitive decline, as lower values were found in PD-sym for the MoCA and hippocampal volume percentile scores.^28^ However, no statistically significant differences were found in cognitive performance or hippocampal volume percentile scores after correction for multiple testing. Differences in cognitive performance might be too subtle to detect with the MoCA at the time of diagnosis A more elaborate neuropsychological assessment and longitudinal follow-up might therefore be needed to reveal differential cognitive decline between both groups.

Motor-symptom severity was higher in PD-sym, which would be in line with the SOC-model. However, PD-sym and PD-asym are contrasted based on the asymmetry of the dopaminergic degeneration. Therefore, this finding would be expected regardless of the SOC-model, as more symmetric dopaminergic degeneration will lead to a more symmetric and severe motor symptom burden. In addition, PD-sym and PD-asym also showed an overrepresentation of PIGD and TD motor subtypes, respectively. Though not explicitly predicted by the SOC-model, the SOC-model and the PIGD/TD-motor classification might be integrated, with PIGD being associated with more symmetric motor involvement in PD. Also, pPD-body and PIGD are both the more malignant subtype in their respective classification.^29,30^ The TD subtype is associated with asymmetric motor involvement, more cerebello-thalamic-cortical dysfunction, and reduced gray matter in the cerebellar vermis.^29,31–33^ Interestingly, the cerebellar vermal lobules VIII-X showed more atrophy in PD-asym, although this difference was not statistically significant after FDR correction. Nonetheless, PIGD and TD at diagnosis might not be mutually exclusive for pPD-brain and pPD-body. Serotonergic projections from the raphe nuclei, located caudally from the SN, have been associated with a dopamine-resistant tremor, whereas cerebello-thalamic-cortical dysfunction might be dopamine-responsive.^34,35^ Longitudinal follow-up of the response to dopaminergic treatment in PD-sym and PD-asym could provide support for this conceptual framework.

Last, PD-sym subjects were older than PD-asym, a difference also found between pPD-body and pPD-brain in the cohort used to describe the brain-first versus body-first dichotomy.^12^ This cannot be attributed to a diagnostic delay, as there was no difference in motor-symptom duration and putaminal SOR at the most affected side between both groups (Table 1, Figure 1). pPD-body is hypothesized to have a longer prodromal period, as the aSyn-pathology needs to cross more synapses before reaching the substantia nigra, compared to suggested pPD-brain sites-of-onset.^11^ In addition, genetic causes of PD are associated with a younger age-of-onset and are thought to be of particular importance in the etiology of pPD-brain. This with the exception of *GBA1* and *SNCA* mutations, which are associated with reduced cardiac innervation and therefore pPD-body.^36,37^ Nonetheless, an older age-of-onset might be part of the clinical subtype of pPD-body and statistical adjustment could lead to overcorrection.

### Brain region volumes and asymmetry

In concordance with the SOC-model, PD-sym is associated with a relative loss of brain volume, in particular of cerebral white matter. However, no differences were found in the BAI of cerebral gray and white matter as measures of overall brain volume, nor in the mean percentile scores and BAIs of brain region volumes after FDR-correction. The lack of identified brain regions with a statistically significant difference after FDR-correction does, however, not directly contradict the SOC-model. The SOC-model describes the propagation of aSyn aggregation.^11^ In the absence of aSyn imaging markers, structural MRI data was used to determine subsequent differential degeneration. However, due to selective neuronal vulnerability, neuronal populations might be invested with aSyn pathology without subsequent cell death.^38^ Also, neurodegeneration in PD is believed to occur in a “dying-back” fashion, starting at the peripheral end of the axon before affecting the cell body.^39,40^ Therefore, neurodegeneration as a consequence of disease progression might not yet be very pronounced on structural MRI in the early phases of the disease.

In line with the SOC-model, the VBM analysis indicates several brain regions with lower gray matter volumes in PD-sym when corrected for TIV, whereas no brain region was identified as smaller in PD-asym. After correction for age, none of the identified regions could be identified at FDR<0.05. Age correction is essential when comparing gray matter structures.^41^ However, this might lead to overcorrection in a direct comparison between both groups, since PD-sym is associated with an older age of onset. The percentile scores provided by cNeuro are normalized for age, based on an internal dataset of control subjects. Therefore, the cNeuro percentile scores potentially provide a more relevant picture. Nonetheless, most identified smaller brain regions in PD-sym were located in the least affected hemisphere, suggesting a relatively intact hemisphere at the side of the smallest dopaminergic deficit in PD-asym. This supports the distribution of degeneration as proposed by the SOC-model, since there is no reason to assume that age-related gray matter atrophy would be restricted to a single hemisphere in the younger PD-asym group.

### Site-of-origin PD-asym

The SOC-model predicts neurodegeneration mainly limited to the hemisphere with the largest dopaminergic deficit in pPD-brain.^11,13^ Limbic structures within the temporal lobe (eg. amygdala and entorhinal area) have been proposed as possible sites-of-origin of pPD-brain.^11,18,19^ The entorhinal area, as well as the transverse temporal gyrus were among the brain regions with the most negative BAI in PD-asym, indicating lower volumes in the most affected hemisphere. On the contrary, the amygdala actually had higher percentile scores in the most affected hemisphere (Supplementary Table 3). Also, the voxel-wise asymmetry analysis of the amygdala and entorhinal cortex showed no difference in asymmetry between both groups. To account for a shared trend of brain regions towards asymmetry, which could be confounded by pre-existent dexterity, a guilt-by-association analysis was performed through correlating BAIs in PD-asym. This analysis yielded a cluster of correlated brain regions including several (limbic) structures within the temporal lobe. Interestingly, the cluster also contained cerebellar white matter, which is more affected in TD subjects,^42^ who were also overrepresented in PD-asym. However, several of the identified brain regions were also strongly correlated in PD-sym. Therefore, the current analysis does not provide support for the amygdala and entorhinal cortex as possible starting point of the aSyn pathology in brain-first PD.

### Limitations

Clearly, the absence of PSG-proven RBD as a marker of pPD-body limits the interpretation of the presented findings. Questionnaire-based assessments of RBD, such as the RBDSQ, are inaccurate for the identification of RBD in *de novo* PD.^23^ In concordance, no differences between PD-sym and PD-asym were found when using the RBDSQ total score and the more sensitive subscore of question 6. Instead, PD subjects were dichotomized in PD-sym and PD-asym, meaning that the clinical and imaging markers cannot be associated directly with pPD-brain or pPD-body. Nonetheless, the SOC-model does dictate a differential clinical picture and spreading pattern related to the dopaminergic asymmetry. If contradicted, this would have been a powerful argument against the SOC-model. However, the clinical comparisons indicate more non-motor symptomatology with a probable neurological substrate below the substantia nigra in PD-sym, which would actually be in line with a relative overrepresentation of pPD-body according to the SOC-model. Moreover, the BAI of several brain regions differed between PD-asym and PD-sym. Although these differences were not statistically significant after correction for multiple testing, together they indicate a trend of neurodegeneration limited to the most affected hemisphere in PD-asym, as suggested by the SOC-model. PSG is an expensive and labor-intensive tool, limiting its usage in large-scale cohort studies. Ideally, clinical markers that are easier to acquire, such as heart rate variability, orthostatic hypotension or a wearable assessment of RBD, should be interrogated for their ability to identify pPD-brain and pPD-body.^43,44^

Second, much like RBD, other non-motor symptoms are notorious for the discrepancy between objective markers and patient-reported complaints.^45,46^ Instead of the questionnaire-based assessment of autonomic dysfunction, objective assessments, such as the colonic transit time or a measurement of orthostatic hypotension, could be used.

Third, as described above, structural MRI data might not yet capture differential neurodegeneration in the early phases of PD. Alternatives more suitable to detect early, axonal neurodegeneration would include tractography, functional MRI and functional PET imaging, though interpretation of functional imaging might be confounded by compensatory mechanisms.^40,47–49^ In addition, differential brain atrophy between PD-sym and PD-asym on structural MRI might become detectable during follow-up.

### Conclusion

Contrasting PD-sym and PD-asym, based on the nigrostriatal dopaminergic deficit, results in the creation of two clinically distinct groups. The higher levels of non-motor symptoms with a probable neurological substrate below the substantia nigra is suggestive of a relative overrepresentation of pPD-body in the PD-sym group. Though overall brain atrophy differed between both groups, no differences between specific brain regions volume and BAI could be found after adjusting for multiple testing. Assessment of the differential degeneration between pPD-body and pPD-brain, as proposed by the SOC-model, might require other imaging modalities (eg. PET-CT, functional MRI and tractography using DTI) and longitudinal structural MRI studies. In addition, clinical markers associated with autonomic failure might provide sufficient discriminatory power to replace PSG-proven RBD as the current gold standard for identifying pPD-body and pPD-brain.

## Methods

### Participants

128 PD subjects from the Dutch Parkinson Cohort study of de novo PD subjects (DUPARC), with complete FDOPA-PET and T1-MRI data, were included in the current study.^20^ Inclusion criteria concerned a movement disorder specialist diagnosis of PD, based on the Movement Disorder Society clinical diagnostic criteria;^1^ and a presynaptic dopaminergic deficiency quantified through 3,4-dihydroxy-6-18F-fluoro-1-phenylalaninie (FDOPA) PET scan imaging. Exclusion criteria were contraindications to MRI, dopaminergic and/or systemic cholinergic medication use, and inability to provide informed consent. All subjects provided written informed consent and the study was approved by the local ethics committee.

Participants were classified as asymmetric (PD-asym) and symmetric (PD-sym) based on the absolute value of the striatal asymmetricity index (SAI) of the striatal-to-occipital ratios (SOR) of the putamen, as measured by FDOPA-PET. SORs of the FDOPA uptake were quantified by dividing the mean activity of the putamen as the volume-of-interest by the mean occipital value. The (absolute value of the) striatal asymmetricity index (SAI) of the nigrostriatal dopaminergic deficit was defined as:^21^

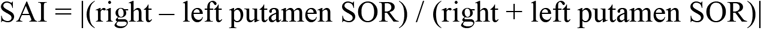

SAIs of both the putamen and caudate were available and showed a strong positive correlated among participants, indicating the dopaminergic deficiency to be on the same side in both the putamen and caudate (Supplementary Figure 1a-c). This was in particular true for PD-asym, in which the most affected side was the same for both the caudate and putamen in 38 participants and differed in only three. Putaminal SAIs were chosen for PD-sym and PD-asym classification as these are more affected at the time of diagnosis and therefore seem to provide the widest range of SAI for participant classification compared to the caudate.^50,51^ Indeed, the absolute SAI of the putamen had a larger range than the SAI of the caudate (Supplementary Figure 1d).

Participants with an SAI within the highest tertile (n=42) were classified as asymmetric (PD-asym) and participants with an SAI in the lowest tertile (n=42) were classified as symmetric (PD-sym). One PD-asym participant had to be removed from the analysis after quality control of MRI data. To balance both groups, one PD-sym participant was removed, making the total number of participants in both groups 41.

### Clinical examination

Motor symptoms were quantified using the Movement Disorders Society Unified Parkinson’s Disease Rating Scale (MDS-UPDRS).^22^ Participants were classified as tremor dominant (TD), postural instability and gait disorder (PIGD) or indeterminate subtypes, based on the ratio of TD and PIGD related items in the MDS-UPDRS part II and part III.^3^ Missing values in the MDS-UPDRS were imputed using the group average of both PD-sym and PD-asym combined, to avoid introducing artificial differences between both groups. Non-motor symptoms were quantified using the Movement Disorders Society Non Motor Symptom Questionnaire (MDS-NMSQ),^52^ the Montreal Cognitive Assessment (MoCA)^53^ and the twelve item Sniffin’ Sticks for olfactory function.^54^ RBD was assessed using the RBD screening questionnaire (RBDSQ)^55^, the scores for question 6 of the RBDSQ were assessed additionally, as question 6 has a higher sensitivity for RBD in *de novo* PD than the RBDSQ total score,^23^

In addition, NMSQ items concerning non-motor symptoms with a probable predominant neurological substrate inferior to the substantia nigra (SN), as a priori judged by a movement disorders specialist (TvL), were grouped together in a below SN subgroup. This concerned the following items.

1: hypersalivation, 3: difficulty swallowing, 4: nausea, 5: constipation, 6: stool incontinence 7: incomplete defecation, 8: urge, 9: nycturia, 19: sexual dysfunction: 20: orthostatic hypotension, 24: lively dreams, 25: RBD, 26: restless legs, 28: hyperhidrosis, 29: diplopia

### Image acquisition

FDOPA-PET was performed after at least six hours of fasting (four hours for diabetic patients). Participants were premedicated with carbidopa 60 minutes before receiving 200MBq of the FDOPA tracer, 90 minutes after which the PET-scan was performed on a Siemens HR+ camera. Subjects received MR imaging using a Siemens Magnetom Prisma 3-Tesla MRI scanner with a 64-channel head coil. Anatomical T1-weighted images were acquired by a sagittal 3-dimensional gradient-echo T1-weighted sequence with 0.9 × 0.9 × 0.9mm acquisition (repetition time 2300 milliseconds, echo time 2.32 milliseconds, inversion time 900 milliseconds).

### Imaging analysis

Quality control of T1-weighted MRI scans was performed using cNeuro automated signal quality control. Furthermore, a visual quality control of the derived structural segmentation in cNeuro was performed by an experienced post-processing specialist (JK) in each individual patient. One scan did not pass quality control, resulting in the exclusion of the participant (part of PD-asym) of all analyses, as well as the exclusion of one PD-sym participant to create a balanced comparison. Images were analyzed using the automated clinical cNeuro software software version 1.11.0 (Combinostics Oy, Tampere, Finland, www.cneuro.com), which quantifies the volumes of predetermined brain regions and several overarching anatomical parameters (eg. cerebral white and gray matter). Results were presented as percentile scores based on an internal reference dataset of 1923 cognitively intact control subjects (18-94 years old, 57% female) normalized for age, sex and head size. Dexterity of brain regions was changed from left/right to most and least affected side according to the dopaminergic deficit. cNeuro was used because it is an automated clinical-grade brain MRI quantification solution, allowing for direct translation of potential findings into clinical practice.

In addition, voxel-wise analyses were applied to compare gray matter volume and asymmetricity between groups. Prior to applying these analyses, MRIs of PD subjects with a larger left hemispheric dopaminergic deficit were flipped using FSL’s fslswapdim function of the FMRIB Software Library (https://fsl.fmrib.ox.ac.uk/fsl/fslwiki/).^56^ Voxel-based morphometry (VBM) and asymmetricity of gray matter structures was performed based on a previously described standardized pipeline using the Statistical Parametric Mapping software version 12 (SPM12).^24,25^

For VBM, each image was manually reoriented to the anterior commissure followed by denoising, bias correction, and tissue segmentation into gray matter, white matter, and cerebrospinal fluid. The tissue segments were then spatially normalized according to the Diffeomorphic Anatomical Registration Through Exponentiated Lie (DARTEL) algebra algorithm.^57^ The resulting maps were then smoothed with an 8 mm FWHM Gausian kernel. An explicit optimal threshold gray matter mask was performed using the SPM Masking toolbox. Anatomical regions were identified and visualized using the xjView toolbox (https://www.alivelearn.net/xjview).

In voxel-wise asymmetry analysis, additional steps were done during the spatial normalization step. The affine-registered gray matter segments were flipped at midline. Then DARTEL spatial normalization was applied for both the original and flipped gray matter segments. The images of each subject were then used to measure asymmetry between the most and least affected hemisphere. The mean asymmetricity index and the volume of two regions regions of interest, the amygdala and the entorhinal cortex, were calculated using smoothed asymmetry index images. The masks of the two regions of interest were obtained from the neuromorphometric atlas provided in SPM12.

### Statistical analysis

Clinical characteristics and cNeuro percentiles were analyzed using R, version 4.0.3. Continuous variables were analyzed with an unpaired t-test or Mann-Whitney U test for parametric and non-parametric continuous variables, respectively. Categorical variables were analyzed by means of a Chi-square test. P-values were adjusted for multiple testing using a false discovery rate (FDR) correction. Brain regions of both hemispheres together were compared using cNeuro percentiles as indicators of volume and relative atrophy. In addition, brain region asymmetricity indices (BAI) were calculated to compare brain region asymmetry between both groups:

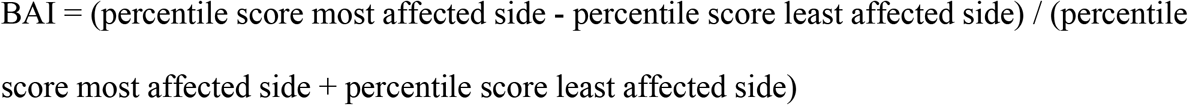

Thereby, a negative BAI represents relatively lower percentile scores of brain region volume at the side of the largest dopaminergic deficit.

BAIs of brain regions in PD-asym were correlated amongst each other, using Pearson correlations, in an exploratory guilt-by-association analysis to identify clusters of brain regions with similar BAI deviations. Euclidean distances of the correlation coefficients were used for hierarchical clustering according to Ward’s method implemented in the R-package heatmap3 (method = “ward.D2”) for visualization.^58^ Pearson correlations of clusters of brain regions identified as highly correlated in PD-asym were also performed in PD-sym to assess whether the found correlations were specific for PD-asym.

Gray matter differences were analyzed by means of a voxel-wise analyses of the smoothed images using SPM12. Differences in gray matter density between PD-sym, and PD-asym were assessed using an unpaired t-test. Results were adjusted in two separate analyses for total intracranial volume (TIV) and for age and TIV, since an age difference between PD-sym and PD-asym might be part of the clinical profiles of pPD-body and pPD-brain. Therefore, correcting for age could lead to overcorrection in the absence of healthy control subjects for the voxel-based analyses. Corrections for multiple comparisons were applied using false discovery rate (FDR) P < 0.05 and a cluster extent threshold of 50 voxels. Additionally, the mean asymmetricity index and volume of two regions of interest, the amygdala and the entorhinal cortex, were calculated and comparisons between PD-asym and PD-sym were done using unpaired t-test.

## Data Availability

Data of the DUPARC study can be requested via the Principal Investigator upon reasonable request.

## Data availability

Data of the DUPARC study can be requested via the Principal Investigator upon reasonable request.^20^

## Funding sources

The DUPARC study was funded by the Weston Brain Institute and personal grants for JMB and MN to cover personnel costs.

We thank all participants in DUPARC study. For their aid in the recruitment of participants, we thank the Parkinson Platform Northern Netherlands (PPNN) Study Group collaborators.* We would like to thank Renée Speijers and Yvonne Nijman for their help in the recruitment and logistics of the study.

*PPNN Study Group

Verwey NA^1^, Van Harten B^1^, Portman AT^2^, Langedijk MJH^2^, Oomes PG^2^, Jansen BJAM^2^, Van Wieren T^2^, Van den Bogaard SJA^3^, Van Steenbergen W^3^, Duyff R^3^, Van Amerongen JP^3^, Fransen PSS^4^, Polman SKL^4^, Zwartbol RT^4^, Van Kesteren ME^4^, Braakhekke JP^4^, Trip J^4^, Koops L^4^, De Langen CJ^4^, De Jong G^4^, Hartono JES^4^, Ybema H^4^, Bartels AL^5^, Reesink FE^5^, Postma AG^6^, Vonk GJH^7^, Oen JMTH^7^, Brinkman MJ^7^, Mondria T^7^, Holscher RS^7^, Van der Meulen AAE^8^, Rutgers AWF^8^, Boekestein WA^9^, Teune LK^9^, Orsel PJL^10^, Hoogendijk JE^10^, Van Laar T^11^.

^1^Department of Neurology, Medisch Centrum Leeuwarden, Leeuwarden, the Netherlands; ^2^Department of Neurology, Treant Zorggroep locations, Stadskanaal, Emmen, Hoogeveen, the Netherlands; ^3^Department of Neurology, Tjongerschans Ziekenhuis Heerenveen, Heerenveen, the Netherlands; ^4^Department of Neurology, Isala, Zwolle, Meppel, the Netherlands; ^5^Department of Neurology, Ommelander Ziekenhuis Groningen, Scheemda, the Netherlands; ^6^Department of Neurology, Nij Smellinghe Ziekenhuis Drachten, Drachten, the Netherlands; ^7^Department of Neurology, Antonius Zorggroep, Sneek, the Netherlands; ^8^Department of Neurology, Martini Ziekenhuis, Groningen, the Netherlands; ^9^Department of Neurology, Wilhelmina Ziekenhuis Assen, Assen, the Netherlands; ^10^Department of Neurology, Sionsberg, Dokkum, the Netherlands; ^11^Department of Neurology, University Medical Center Groningen, Groningen, the Netherlands.

## Authors’ contributions

JMB contributed to the study conception, participant inclusion, data collection, data analysis, writing the first draft of the manuscript and later reviewing of the manuscript. MN contributed to data analysis, writing the first draft of the manuscript and reviewing of the manuscript. JK contributed to data analysis and reviewing of the manuscript. ACS contributed to data collection and reviewing of the manuscript. SvdZ contributed to the study conception, participant inclusion, data collection and reviewing of the manuscript. RJHB contributed to data analysis and reviewing of the manuscript. TvL contributed to the study conception, participant inclusion, data collection and reviewing of the manuscript.

JMB and MN should be considered joint first authors. All authors approve the completed version of the manuscript and are accountable for all aspects of the work in ensuring that questions related to the accuracy or integrity of any part of the work are appropriately investigated and resolved.

