## Supplementary Materials for "Striatal Dopaminergic Asymmetry as a marker of Brain-First and Body-First Subtypes in de novo Parkinson’s Disease"

### Contents


- Supplementary figure 1: Striatal-occipital-binding ratios asymmetric and symmetric Parkinson’s disease (p. 2)
- Supplementary table 1: Percentile scores total brain region volumes cNeuro (p. 3)
- Supplementary table 2: Voxel-based-morphometry analysis adjusted for total intracranial volume (p. 6)
- Supplementary table 3: Brain region asymmetry indices (p. 7)
- Supplementary figure 2: Voxel-wise region-of-interest analyses amygdala and entorhinal cortex (p. 10)
- Supplementary table 4: Guilt-by-association correlation analysis brain region asymmetry indices asymmetric Parkinson’s disease (p. 11)

**Supplementary Figure 1. Comparisons between striatal asymmetry indices (SAIs) of the putamen and caudate.**

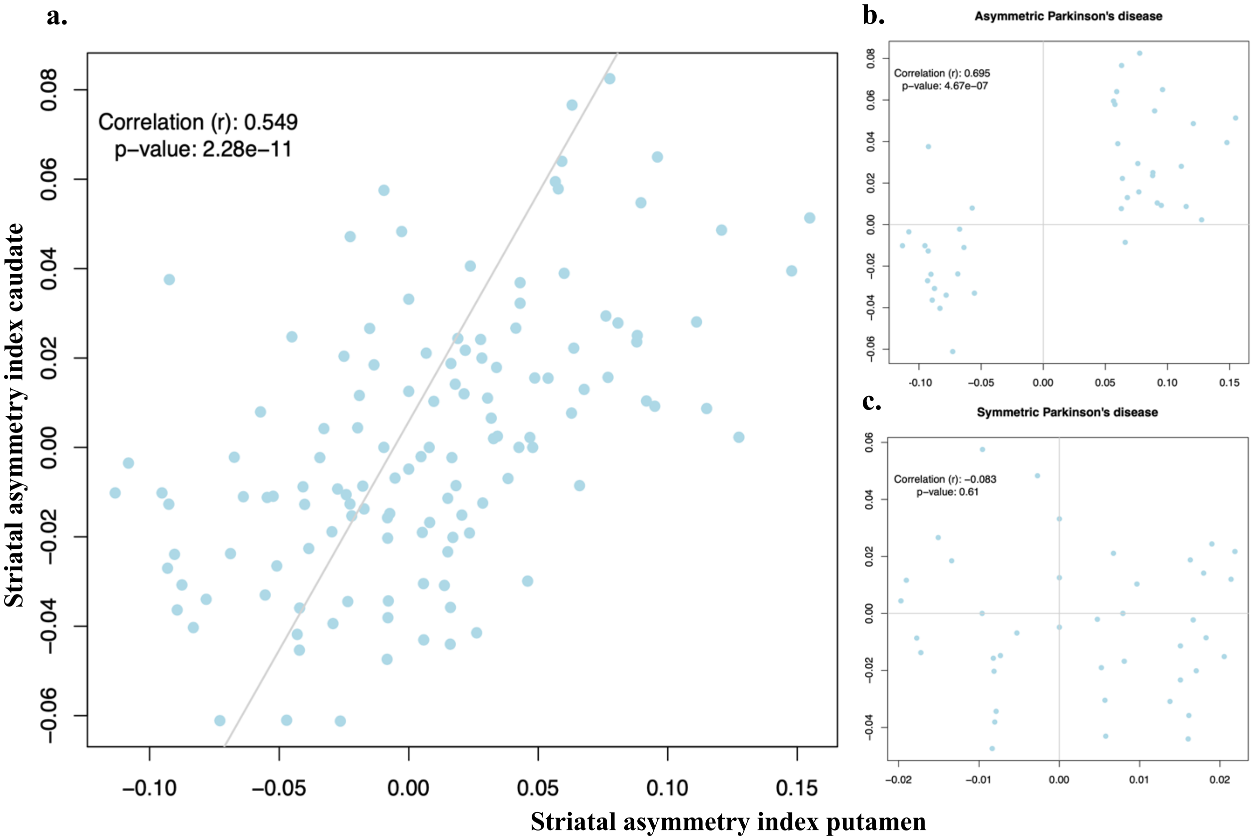


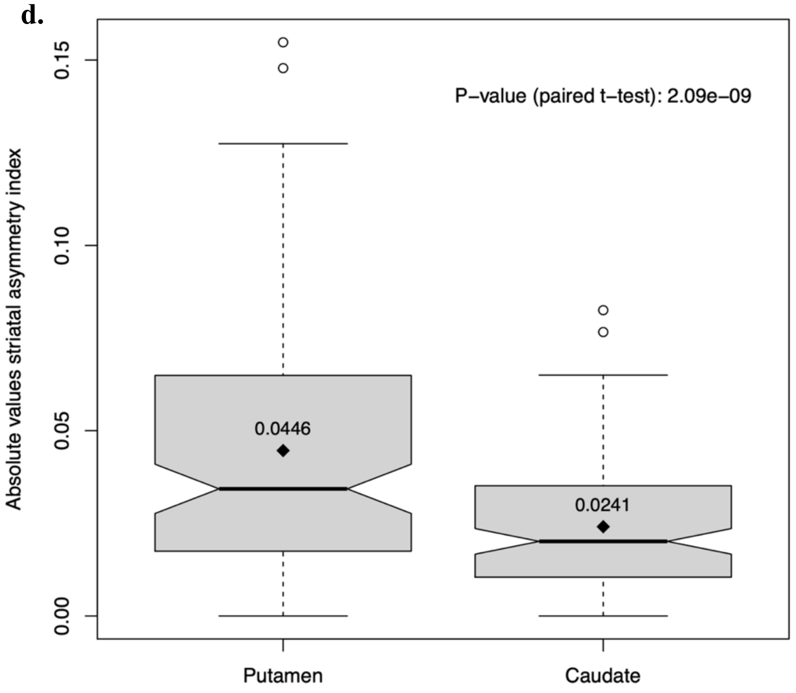


Correlations were performed in (a) all participants (n=127, after exclusion of one participant with an anatomical anomaly of the caudate, which influenced the caudate binding ratio); (b) participants classified as asymmetric Parkinson's disease (PD-asym) and (c) participants classified as symmetric Parkinson's disease (PD-sym). A strong positive correlation was found between the SAIs of the caudate and putamen, mainly driven by PD-asym and confounded by PD-sym. (d) The range in absolute putaminal SAIs is almost twice as large as the range of the caudate SAIs (n=127), resulting in a statistically significant difference in the mean striatal asymmetry indices between both areas (paired t-test).

**Supplementary table 1.** Percentile scores total brain region volumes cNeuro

| | mean_sym | sd_sym | mean_asym | sd_asym | ttest_nominal_p | ttest_fdr |
|---|---|---|---|---|---|---|
| **Brain_stem** | 51,14573171 | 32,38216616 | 57,53812195 | 29,04558469 | 0,349597005 | 0,812267118 |
| **Cerebellar_vermal_lobules_I,V** | 45,07582927 | 29,93014708 | 46,65497561 | 30,26938468 | 0,812848409 | 0,885506419 |
| **Cerebellar_vermal_lobules_VI,VII** | 56,40419512 | 31,77468874 | 51,74470732 | 31,2196698 | 0,504932758 | 0,812267118 |
| **Cerebellar_vermal_lobules_VIII,X** | 65,04582927 | 27,22274679 | 45,02834146 | 34,43447393 | 0,004605209 | 0,290128145 |
| **Hippocampus** | 41,47056098 | 28,25810821 | 54,67009756 | 27,51266005 | 0,035154968 | 0,592734792 |
| **Accumbens_Area** | 38,70687805 | 25,81276066 | 41,88417073 | 27,83466565 | 0,593503196 | 0,824805723 |
| **Amygdala** | 31,9805122 | 21,47000824 | 41,33804878 | 28,06245817 | 0,094084888 | 0,592734792 |
| **Caudate** | 38,84756098 | 29,09926461 | 41,7117561 | 31,99159653 | 0,672656297 | 0,87006562 |
| **Cerebellum_Exterior** | 68,43785366 | 26,57980483 | 55,80843902 | 33,78435277 | 0,06378195 | 0,592734792 |
| **Cerebellum_White_Matter** | 68,57931707 | 26,93074394 | 75,13482927 | 20,38928618 | 0,21788344 | 0,812267118 |
| **Pallidum** | 52,18839024 | 30,91229826 | 65,76858537 | 28,25214327 | 0,041092478 | 0,592734792 |
| **Putamen** | 30,51326829 | 28,25583608 | 28,54317073 | 27,48315851 | 0,749778835 | 0,874741974 |
| **Thalamus_proper** | 28,46190244 | 22,71246651 | 23,08368293 | 25,01800226 | 0,311226245 | 0,812267118 |
| **Ventral_DC** | 48,44902439 | 30,57052286 | 56,00660976 | 28,28389489 | 0,248735761 | 0,812267118 |
| **Anterior_cingulate_gyrus** | 46,37692683 | 29,49513844 | 50,12885366 | 36,44955388 | 0,609868319 | 0,824805723 |
| **Anterior_insula** | 37,97585366 | 31,70650265 | 42,8255122 | 25,75591965 | 0,449473617 | 0,812267118 |
| **Anterior_orbital_gyrus** | 34,17787805 | 24,19358231 | 43,12290244 | 31,07890891 | 0,150027181 | 0,717638206 |
| **Angular_gyrus** | 37,43860976 | 31,22025153 | 44,81007317 | 32,41187397 | 0,297415771 | 0,812267118 |
| **Calcarine_cortex** | 54,33980488 | 33,41127069 | 58,7405122 | 30,70160681 | 0,536372847 | 0,812267118 |
| **Central_operculum** | 44,53060976 | 26,6110936 | 45,81968293 | 33,87756934 | 0,848566915 | 0,885506419 |
| **Cuneus** | 62,85795122 | 32,36458479 | 66,9727561 | 30,36606562 | 0,554404541 | 0,812267118 |
| **Entorhinal_area** | 45,4645122 | 30,35902874 | 50,3764878 | 27,79329295 | 0,44704831 | 0,812267118 |
| **Frontal_operculum** | 39,64765854 | 28,47655449 | 47,08697561 | 35,11044696 | 0,295322722 | 0,812267118 |
| **Frontal_pole** | 34,23360976 | 28,02767065 | 35,18343902 | 24,85821768 | 0,871450761 | 0,885506419 |
| **Fusiform_gyrus** | 59,79021951 | 23,56544224 | 63,58065854 | 31,53047555 | 0,539406213 | 0,812267118 |
| **Gyrus_rectus** | 31,214 | 26,39027048 | 35,14770732 | 27,23343738 | 0,508474487 | 0,812267118 |
| **Inferior_occipital_gyrus** | 42,14553659 | 29,4562408 | 41,4804878 | 27,16059744 | 0,915626726 | 0,915626726 |
| **Inferior_temporal_gyrus** | 48,59570732 | 29,66564997 | 51,18636585 | 33,27542804 | 0,710808622 | 0,874741974 |
| **Lingual_gyrus** | 63,4627561 | 29,14075344 | 65,64802439 | 27,98585239 | 0,730008768 | 0,874741974 |

| | | | | | | |
|---|---|---|---|---|---|---|
| **Lateral_orbital_gyrus** | 32,08063415 | 25,7157385 | 41,99536585 | 32,87274798 | 0,132404259 | 0,695122359 |
| **Middle_cingulate_gyrus** | 52,75209756 | 32,59395549 | 47,14258537 | 34,818612 | 0,453607742 | 0,812267118 |
| **Medial_frontal_cortex** | 42,39160976 | 26,6843482 | 47,18914634 | 29,48512171 | 0,442130513 | 0,812267118 |
| **Middle_frontal_gyrus** | 29,72370732 | 26,65549338 | 38,53917073 | 32,73017799 | 0,185088289 | 0,777370814 |
| **Middle_occipital_gyrus** | 37,26387805 | 30,64310891 | 48,96214634 | 28,47760131 | 0,077161256 | 0,592734792 |
| **Medial_orbital_gyrus** | 37,25458537 | 27,28933021 | 32,68112195 | 26,36150123 | 0,442503264 | 0,812267118 |
| **Postcentral_gyrus_medial_segment** | 60,75329268 | 32,00005074 | 48,25058537 | 34,47900685 | 0,092686111 | 0,592734792 |
| **Precentral_gyrus_medial_segment** | 44,94380488 | 31,27664308 | 55,77021951 | 29,21055592 | 0,109218316 | 0,625523083 |
| **Superior_frontal_gyrus_medial_segment** | 34,44173171 | 29,44215159 | 33,12126829 | 24,26432159 | 0,825199499 | 0,885506419 |
| **Middle_temporal_gyrus** | 43,55587805 | 31,82968735 | 45,85360976 | 37,53766438 | 0,765780774 | 0,877167069 |
| **Occipital_pole** | 16,01185366 | 18,42027728 | 20,91817073 | 24,29225923 | 0,306113467 | 0,812267118 |
| **Occipital_fusiform_gyrus** | 60,61270732 | 27,37547457 | 64,68465854 | 27,02134319 | 0,499829678 | 0,812267118 |
| **Opercular_part_of_the_inferior_frontal_gyrus** | 39,41173171 | 32,9580135 | 43,07707317 | 32,83833766 | 0,615331253 | 0,824805723 |
| **Orbital_part_of_the_inferior_frontal_gyrus** | 46,52273171 | 26,67856172 | 47,58243902 | 31,91399116 | 0,870843384 | 0,885506419 |
| **Posterior_cingulate_gyrus** | 60,69097561 | 29,72701958 | 59,56204878 | 29,29887755 | 0,862940913 | 0,885506419 |
| **Precuneus** | 51,6614878 | 26,67376326 | 58,67487805 | 32,25744869 | 0,28666661 | 0,812267118 |
| **Parahippocampal_gyrus** | 56,35141463 | 27,14513738 | 51,53058537 | 28,59424361 | 0,435989287 | 0,812267118 |
| **Posterior_insula** | 34,20980488 | 29,02142619 | 46,39004878 | 33,07582307 | 0,080196146 | 0,592734792 |
| **Parietal_operculum** | 52,76480488 | 33,77443871 | 55,01807317 | 28,89073605 | 0,746334376 | 0,874741974 |
| **Postcentral_gyrus** | 45,9712439 | 31,13681679 | 36,74380488 | 27,58923013 | 0,159475157 | 0,717638206 |
| **Posterior_orbital_gyrus** | 54,33614634 | 26,02186857 | 48,19009756 | 32,62635516 | 0,34866008 | 0,812267118 |
| **Planum_polare** | 37,50970732 | 29,04922739 | 45,30221951 | 28,86632578 | 0,226656128 | 0,812267118 |
| **Precentral_gyrus** | 42,41441463 | 30,8501992 | 36,12941463 | 29,1419784 | 0,345846412 | 0,812267118 |
| **Planum_temporale** | 46,0817561 | 33,60522753 | 42,16268293 | 32,39839636 | 0,592355501 | 0,824805723 |
| **Subcallosal_area** | 55,48095122 | 31,42339393 | 52,94231707 | 27,02944146 | 0,695996374 | 0,874741974 |
| **Superior_frontal_gyrus** | 37,09392683 | 30,2288743 | 41,50031707 | 30,54215027 | 0,513335581 | 0,812267118 |
| **Supplementary_motor_cortex** | 36,03104878 | 30,72685617 | 47,81617073 | 27,82453448 | 0,072471791 | 0,592734792 |
| **Supramarginal_gyrus** | 50,34080488 | 29,52759965 | 34,78178049 | 30,51887727 | 0,021450696 | 0,592734792 |
| **Superior_occipital_gyrus** | 36,33336585 | 30,10848608 | 34,76295122 | 31,27994707 | 0,817433875 | 0,885506419 |
| **Superior_parietal_lobule** | 43,94609756 | 30,35449781 | 40,16063415 | 25,96441492 | 0,545733219 | 0,812267118 |
| **Superior_temporal_gyrus** | 43,80673171 | 33,42528657 | 37,26563415 | 29,81606519 | 0,352595726 | 0,812267118 |
| **Temporal_pole** | 38,3105122 | 27,05519034 | 43,477 | 32,40252988 | 0,435609129 | 0,812267118 |

| | | | | | | |
|---|---|---|---|---|---|---|
| **Triangular_part_of_the_inferior_frontal_gyrus** | 36,12941463 | 30,57170138 | 42,21831707 | 29,45129561 | 0,361151209 | 0,812267118 |
| **Transverse_temporal_gyrus** | 52,80760976 | 34,16294636 | 49,76521951 | 31,62005116 | 0,676717704 | 0,87006562 |
| **Cumulative measures:** | | | | | | |
| **Brain_tissue_all_regions** | 53,8755122 | 25,66648563 | 65,17909756 | 19,96826219 | 0,029020707 | |
| **Cerebral_gray_matter** | 30,25631707 | 25,44125681 | 35,55970732 | 30,43021649 | 0,394556332 | |
| **Cerebral_white_matter** | 53,56073171 | 25,04484618 | 70,2014878 | 24,79036486 | 0,003355222 | |
| **Cerebrospinal_fluid** | 65,45687805 | 25,36019274 | 58,10736585 | 28,77357681 | 0,223487447 | |

**Supplementary Table 2. Differences in gray matter density between symmetric and asymmetric PD based on voxel-based morphometry**

| No. | Voxel Size | T | Z score | MNI Coordinates x | y | z | Location of local maxima |
|---|---|---|---|---|---|---|---|
| **A. Symmetric PD < asymmetric PD most-affected hemisphere** | | | | | | | |
| 1 | 7559 | 4.69 | 4.39 | -60 | -44 | -15 | Temporal Inferior |
| 2 | 3858 | 4.55 | 4.28 | -8 | -20 | 59 | Paracentral lobule |
| 3 | 769 | 4.17 | 3.95 | -26 | -6 | -9 | Amygdala |
| 4 | 346 | 3.92 | 3.74 | -20 | -62 | 56 | Parietal Superior |
| 5 | 1192 | 3.87 | 3.69 | -51 | 27 | 27 | Frontal Inferior pars Triangularis |
| 6 | 132 | 3.7 | 3.55 | -14 | -59 | 72 | Precuneus |
| 7 | 514 | 3.69 | 3.53 | -57 | 26 | 2 | Frontal Inferior pars Triangularis |
| 8 | 131 | 3.63 | 3.48 | -9 | -78 | 45 | Precuneus |
| 9 | 222 | 3.56 | 3.41 | -17 | 62 | 24 | Frontal Superior |
| 10 | 294 | 3.52 | 3.38 | -35 | -11 | 6 | Insula |
| 11 | 628 | 3.45 | 3.32 | -9 | 15 | 6 | Caudate |
| 12 | 157 | 3.36 | 3.24 | -17 | -33 | 2 | Hippocampus |
| 13 | 108 | 3.29 | 3.17 | -30 | 5 | -32 | Superior Temporal Pole |
| 14 | 66 | 3.13 | 3.03 | -44 | 3 | 3 | Insula |
| 15 | 64 | 3.04 | 2.94 | -8 | 26 | -20 | Gyrus Rectus |
| 16 | 54 | 2.98 | 2.89 | -12 | -74 | 21 | Cuneus |
| 17 | 50 | 2.97 | 2.88 | -3 | -48 | 63 | Precuneus |
| **B. Symmetric PD < asymmetric PD least-affected hemisphere** | | | | | | | |
| **No.** | **Voxel Size** | **T** | **Z score** | **MNI Coordinates x** | **y** | **z** | **Location of local maxima** |
| 18 | 5191 | 4.44 | 4.18 | 6 | -74 | 5 | Calcarine |
| 19 | 1909 | 4.03 | 3.83 | 6 | 42 | -11 | Medial Orbitofrontal cortex |
| 20 | 280 | 3.43 | 3.3 | 6 | 36 | 17 | Cingulum Anterior |
| 21 | 328 | 3.44 | 3.31 | 11 | 15 | 8 | Caudate |
| 22 | 536 | 4.53 | 4.26 | 23 | -87 | 39 | Occipital Superior |
| 23 | 171 | 3.32 | 3.2 | 23 | -3 | -9 | Amygdala |
| 24 | 160 | 3.36 | 3.24 | 32 | -15 | -12 | Hippocampus |
| 25 | 989 | 4.18 | 3.96 | 36 | -8 | 11 | Insula |
| 26 | 544 | 3.57 | 3.42 | 44 | -68 | -8 | Temporal Inferior |
| 27 | 518 | 3.42 | 3.29 | 50 | -54 | 44 | Parietal Inferior |
| 28 | 1865 | 3.92 | 3.74 | 53 | 35 | 17 | Frontal Inferior pars Triangularis |

Brain areas with a statistically significantly lower gray matter density, corrected for total intracranial volume, in symmetric PD compared to asymmetric PD in the most-affected hemisphere (1A) and least-affected hemisphere (1B) according to the dopaminergic deficit quantified by FDOPA-PET. No brain areas were identified with a lower gray matter density in asymmetric PD compared to symmetric PD. Statistically significant results are reported at false discovery rate $p$<0.05 with an extended threshold $k$ = 50. MNI, Montreal Neurological Institute

**Supplementary table 3.** Brain regions asymmetry indices (BAIs) cNeuro

| | mean_sym | sd_sym | mean_asym | sd_asym | ttest_nominal_p | ttest_fdr |
|---|---|---|---|---|---|---|
| **Accumbens_area** | -0,000300076 | 0,471111961 | 0,021765231 | 0,486525143 | 0,8352743 | 0,997988557 |
| **Amygdala** | -0,047989773 | 0,455927412 | 0,018087851 | 0,34725844 | 0,462672009 | 0,997988557 |
| **Caudate** | 0,049706797 | 0,254382537 | 0,025404493 | 0,283202675 | 0,683809783 | 0,997988557 |
| **Cerebellum_exterior** | 0,038297338 | 0,087007561 | -0,010684394 | 0,203545217 | 0,16225517 | 0,9440567 |
| **Cerebellum_white_matter** | -0,005715808 | 0,217510101 | -0,016939423 | 0,132823723 | 0,778834934 | 0,997988557 |
| **Hippocampus** | -0,065620373 | 0,308985095 | -0,042161167 | 0,215417269 | 0,691233249 | 0,997988557 |
| **Pallidum** | -0,036426298 | 0,315824115 | -0,011546371 | 0,202279452 | 0,672344306 | 0,997988557 |
| **Putamen** | 0,09238955 | 0,329963015 | 0,051654879 | 0,266713673 | 0,540534308 | 0,997988557 |
| **Thalamus_proper** | 0,003256674 | 0,392459015 | -0,099579616 | 0,259504524 | 0,166110502 | 0,9440567 |
| **Ventral_diencephalon** | -0,014687948 | 0,133255536 | -0,020870966 | 0,155897309 | 0,847426891 | 0,997988557 |
| **Anterior_cingulate_gyrus** | -0,165172755 | 0,449155568 | -0,016379546 | 0,482274461 | 0,152199113 | 0,9440567 |
| **Anterior_insula** | -0,032957969 | 0,410390177 | 0,049916546 | 0,314129819 | 0,307828407 | 0,997988557 |
| **Anterior_orbital_gyrus** | 0,056013697 | 0,570006668 | -0,03171591 | 0,47466704 | 0,451164142 | 0,997988557 |
| **Angular_gyrus** | 0,135121619 | 0,465674723 | 0,075612159 | 0,521913902 | 0,587446098 | 0,997988557 |
| **Calcarine_cortex** | -0,057452101 | 0,33389825 | 0,01245156 | 0,340577158 | 0,350829879 | 0,997988557 |
| **Central_operculum** | 0,048451703 | 0,510194003 | 0,092721959 | 0,524703813 | 0,699542804 | 0,997988557 |
| **Cuneus** | -0,064545581 | 0,397434174 | 0,017100599 | 0,398253225 | 0,355590798 | 0,997988557 |
| **Entorhinal_area** | -0,009256371 | 0,451288892 | -0,13232004 | 0,356451477 | 0,174654563 | 0,9440567 |
| **Frontal_operculum** | -0,096204744 | 0,461338501 | -0,085620513 | 0,456620701 | 0,917106286 | 0,997988557 |
| **Frontal_pole** | -0,071065062 | 0,571466539 | -0,052957237 | 0,454563065 | 0,874257773 | 0,997988557 |
| **Fusiform_gyrus** | 0,016055412 | 0,376147013 | 0,011996748 | 0,348047594 | 0,95968243 | 0,997988557 |
| **Gyrus_rectus** | 0,037312916 | 0,506352282 | -0,024564481 | 0,467624617 | 0,567022535 | 0,997988557 |
| **Inferior_occipital_gyrus** | -0,042848431 | 0,564596998 | -0,021262959 | 0,452976778 | 0,849076141 | 0,997988557 |
| **Inferior_temporal_gyrus** | -0,027163683 | 0,409618682 | 0,01051513 | 0,400677191 | 0,674847881 | 0,997988557 |
| **Lingual_gyrus** | -0,034553883 | 0,277964188 | -0,008499972 | 0,297970013 | 0,683347353 | 0,997988557 |
| **Lateral_orbital_gyrus** | -0,104483253 | 0,498442986 | 0,032502261 | 0,50925098 | 0,221963139 | 0,994943165 |
| **Middle_cingulate_gyrus** | 0,063429343 | 0,43183458 | 0,011298344 | 0,505092796 | 0,61686123 | 0,997988557 |
| **Medial_frontal_cortex** | 0,033674424 | 0,509727175 | 0,041832586 | 0,51441065 | 0,942675813 | 0,997988557 |
| **Middle_frontal_gyrus** | 0,173383418 | 0,491972427 | -0,031658065 | 0,538247253 | 0,075589006 | 0,919572484 |

| | | | | | | |
|---|---|---|---|---|---|---|
| **Middle_occipital_gyrus** | 0,076487463 | 0,516178028 | -0,053035293 | 0,457073552 | 0,232615821 | 0,994943165 |
| **Medial_orbital_gyrus** | -0,004114349 | 0,44958087 | -0,034805212 | 0,478699659 | 0,765534125 | 0,997988557 |
| **Postcentral_gyrus_medial_segment** | -0,070506822 | 0,423603558 | 0,03121376 | 0,472833104 | 0,308026543 | 0,997988557 |
| **Precentral_gyrus_medial_segment** | 0,018670448 | 0,403345441 | -0,105482313 | 0,41996872 | 0,176010571 | 0,9440567 |
| **Superior_frontal_gyrus_medial_segment** | 0,021229941 | 0,546151 | -0,043794084 | 0,537197865 | 0,588299008 | 0,997988557 |
| **Middle_temporal_gyrus** | 0,128673951 | 0,473391412 | -0,11182426 | 0,406080639 | 0,015728204 | 0,463982023 |
| **Occipital_pole** | 0,054479703 | 0,582807482 | 0,047096754 | 0,601105375 | 0,95511376 | 0,997988557 |
| **Occipital_fusiform_gyrus** | -0,074365875 | 0,377291418 | 0,073176055 | 0,370852035 | 0,077929872 | 0,919572484 |
| **Opercular_part_of_the_inferior_frontal_gyrus** | 0,053766507 | 0,459456763 | 0,053488159 | 0,534366456 | 0,997988557 | 0,997988557 |
| **Orbital_part_of_the_inferior_frontal_gyrus** | 0,079181445 | 0,540164119 | 0,118316737 | 0,570118513 | 0,750508594 | 0,997988557 |
| **Posterior_cingulate_gyrus** | 0,044377145 | 0,285162263 | -0,100730317 | 0,305623663 | 0,029063249 | 0,571577238 |
| **Precuneus** | 0,062632141 | 0,331814169 | 0,147026909 | 0,307880154 | 0,236088209 | 0,994943165 |
| **Parahippocampal_gyrus** | 0,022920581 | 0,284094774 | -0,027820599 | 0,349197501 | 0,472645724 | 0,997988557 |
| **Posterior_insula** | -0,030084655 | 0,482513628 | -0,026260012 | 0,462268961 | 0,970856057 | 0,997988557 |
| **Parietal_operculum** | -0,009301461 | 0,309698724 | -0,047904662 | 0,417127294 | 0,63563546 | 0,997988557 |
| **Postcentral_gyrus** | 0,024084104 | 0,485735701 | -0,005968864 | 0,54713448 | 0,793223729 | 0,997988557 |
| **Posterior_orbital_gyrus** | 0,057639441 | 0,419746906 | -0,008866889 | 0,399160531 | 0,464380779 | 0,997988557 |
| **Planum_polare** | 0,055285686 | 0,523909466 | 0,040677803 | 0,427166469 | 0,890309332 | 0,997988557 |
| **Precentral_gyrus** | 0,022637204 | 0,419437208 | -0,053001813 | 0,448364465 | 0,432544998 | 0,997988557 |
| **Planum_temporale** | -0,086544559 | 0,492506538 | 0,022642484 | 0,508657056 | 0,326400826 | 0,997988557 |
| **Subcallosal_area** | -0,008715797 | 0,225968143 | -0,009318935 | 0,322631321 | 0,992204445 | 0,997988557 |
| **Superior_frontal_gyrus** | -0,128217693 | 0,438954069 | -0,169262864 | 0,41136502 | 0,663382866 | 0,997988557 |
| **Supplementary_motor_cortex** | -0,058316457 | 0,527370603 | -0,058838575 | 0,393256016 | 0,99595887 | 0,997988557 |
| **Supramarginal_gyrus** | -0,046383086 | 0,423482281 | 0,000574093 | 0,491165452 | 0,644200819 | 0,997988557 |
| **Superior_occipital_gyrus** | 0,003683075 | 0,524535942 | 0,000907675 | 0,578237289 | 0,981896434 | 0,997988557 |
| **Superior_parietal_Lobule** | 0,02591888 | 0,350864941 | -0,111012393 | 0,414973043 | 0,110692705 | 0,9440567 |
| **Superior_temporal_gyrus** | 0,023733344 | 0,527707231 | 0,015195271 | 0,502916816 | 0,940405163 | 0,997988557 |
| **Temporal_pole** | 0,037592514 | 0,444269909 | -0,013982124 | 0,43664535 | 0,5974819 | 0,997988557 |
| **Triangular_part_of_the_inferior_frontal_gyrus** | 0,032947044 | 0,618411068 | 0,066293294 | 0,438781819 | 0,779063076 | 0,997988557 |
| **Transverse_temporal_gyrus** | 0,142823575 | 0,448378181 | -0,160462708 | 0,44495757 | 0,002886129 | 0,170281609 |
| **Cumulative measures:** | | | | | | |
| **Cerebral_white_matter** | -0,01802637 | 0,132663134 | -0,019861658 | 0,110290767 | 0,945868333 | |

| | | | | | |
|---|---|---|---|---|---|
| **Cerebral_gray_matter** | 0,044174777 | 0,302576166 | -0,069704955 | 0,28968694 | 0,085579816 |

**Supplementary figure 2. Voxel-wise region-of-interest-based analysis of the volume and the asymmetric index of amygdala and entorhinal cortex.**

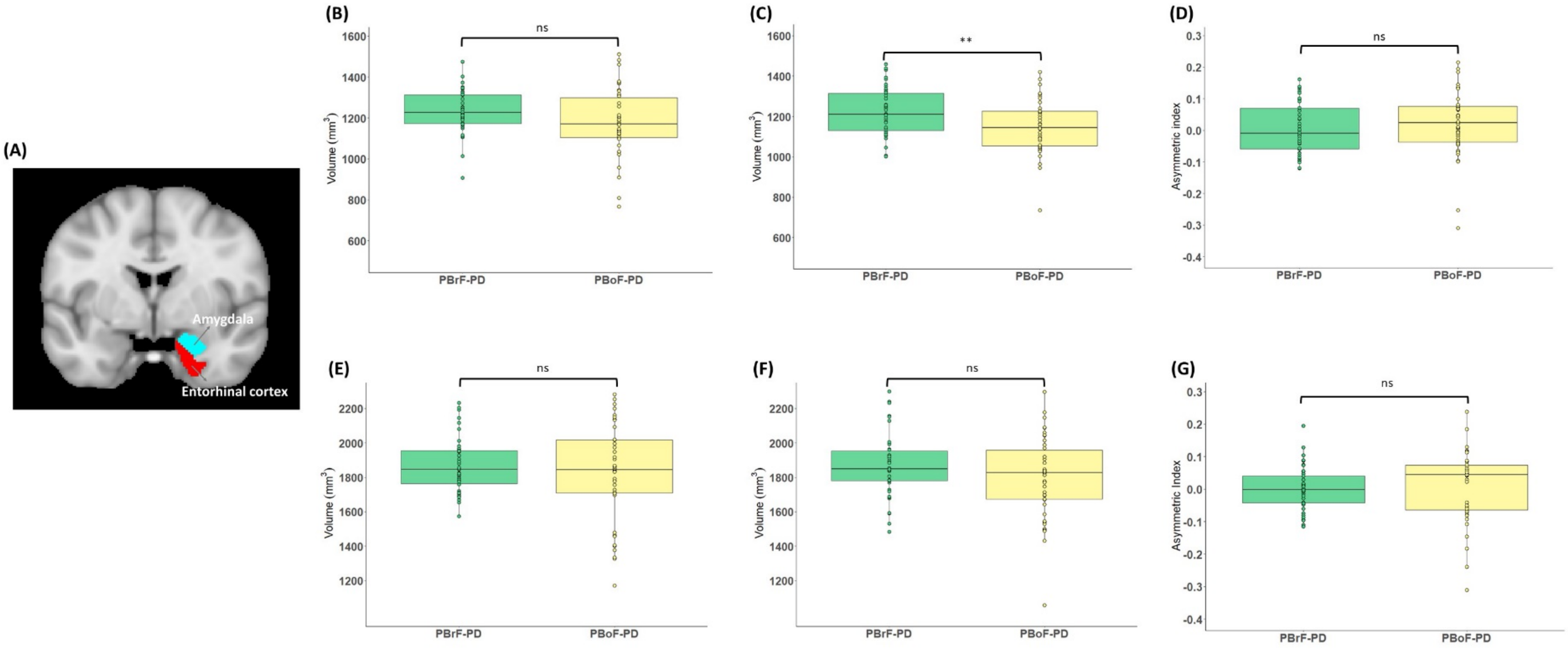


The masks of the two regions are depicted in panel A. The volume comparisons of the amygdala between asymmetric, enriched with probable-brain-first PD (PBrF-PD) and symmetric, enriched with probable-body-first-PD (PBoF-PD) for the brain hemisphere with the most-dopaminergic-deficit side and the least-dopaminergic-deficit side are shown in panel B and C, respectively. Similar volumetric comparisons for entorhinal cortex are visualized in panel E and F. Finally, between-group-comparisons of the asymmetry index of amygdala and entorhinal cortex are presented in panel D and G, respectively. PBrF-PD group had significantly higher amygdala volume in the least-dopaminergic-deficit hemisphere compared to PBoF-PD. ($^{ns}P > 0.05$; $^{**}P < 0.01$).

**Supplementary table 4.** Guilt-by-association correlation analysis brain region asymmetry indices asymmetric Parkinson's disease

**Supplementary table 4A. Brain regions per cluster (displayed from top right to bottom left in heatmap figure 4a)**

| **Cluster 1** | **Cluster 2** | **Cluster 3** |
|---|---|---|
| Amygdala | Thalamus proper | Accumbens area |
| Cerebellum white matter | Ventral diencephalon | Caudate |
| Hippocampus | Anterior insula | Cerebellum exterior |
| Putamen | Angular gyrus | Pallidum |
| Anterior orbital gyrus | Calcarine cortex | Anterior cingulate gyrus |
| Central operculum | Inferior occipital gyrus | Cuneus |
| Entorhinal area | Lateral orbital gyrus | Frontal operculum |
| Inferior temporal gyrus | Middle frontal gyrus | Frontal pole |
| Lingual gyrus | Medial orbital gyrus | Fusiform gyrus |
| Precuneus | Precentral gyrus medial segment | Gyrus rectus |
| Planum temporale | Superior frontal gyrus medial segment | Middle cingulate gyrus |
| Superior frontal gyrus | Occipital pole | Medial frontal cortex |
| Superior temporal gyrus | Opercular part of the inferior frontal gyrus | Middle occipital gyrus |
| Temporal pole | Posterior cingulate gyrus | Postcentral gyrus medial segment |
| Triangular part inferior frontal gyrus | Parietal operculum | Middle temporal gyrus |
| | Planum polare | Occipital fusiform gyrus |
| | Supplementary motor cortex | Orbital part of the inferior frontal gyrus |
| | Superior parietal Lobule | Parahippocampal gyrus |
| | | Posterior insula |
| | | Postcentral gyrus |
| | | Posterior orbital gyrus |
| | | Precentral gyrus |
| | | Subcallosal area |
| | | Supramarginal gyrus |
| | | Superior occipital gyrus |
| | | Transverse temporal gyrus |

**Supplementary table 4B. Pearson correlation coefficients in PD-asym of brain regions identified in cluster 1**

| | Amygdala | Cerebellum white matter | Hippocampus | Putamen | Anterior orbital gyrus | Central operculum | Entorhinal area | Inferior temporal gyrus | Lingual gyrus | Precuneus | Planum temporale | Superior frontal gyrus | Superior temporal gyrus | Temporal pole | Triangular part inferior frontal gyrus |
|---|---|---|---|---|---|---|---|---|---|---|---|---|---|---|---|
| **Amygdala** | | 0,413 | 0,109 | 0,043 | 0,123 | 0,332 | 0,297 | -0,012 | 0,152 | -0,016 | 0,166 | 0,023 | 0,165 | 0,498 | 0,209 |
| **Cerebellum white matter** | 0,413 | | -0,075 | -0,192 | -0,190 | 0,193 | 0,208 | 0,010 | 0,094 | 0,142 | 0,138 | -0,197 | 0,211 | 0,136 | 0,093 |
| **Hippocampus** | 0,109 | -0,075 | | 0,139 | 0,094 | -0,047 | 0,220 | 0,048 | 0,130 | 0,038 | 0,037 | 0,331 | 0,452 | 0,079 | 0,177 |
| **Putamen** | 0,043 | -0,192 | 0,139 | | 0,421 | 0,294 | -0,094 | 0,117 | 0,411 | 0,197 | -0,099 | 0,039 | -0,302 | 0,211 | -0,161 |
| **Anterior orbital gyrus** | 0,123 | -0,190 | 0,094 | 0,421 | | 0,144 | -0,231 | 0,370 | 0,295 | 0,098 | -0,342 | 0,048 | -0,157 | 0,149 | 0,016 |
| **Central operculum** | 0,332 | 0,193 | -0,047 | 0,294 | 0,144 | | 0,145 | -0,140 | 0,304 | 0,219 | 0,319 | 0,037 | 0,165 | 0,443 | 0,104 |
| **Entorhinal area** | 0,297 | 0,208 | 0,220 | -0,094 | -0,231 | 0,145 | | -0,018 | -0,160 | 0,099 | 0,260 | 0,383 | 0,346 | 0,269 | 0,077 |
| **Inferior temporal gyrus** | -0,012 | 0,010 | 0,048 | 0,117 | 0,370 | -0,140 | -0,018 | | 0,364 | 0,391 | -0,156 | 0,125 | 0,056 | -0,124 | -0,107 |
| **Lingual gyrus** | 0,152 | 0,094 | 0,130 | 0,411 | 0,295 | 0,304 | -0,160 | 0,364 | | 0,376 | 0,036 | 0,030 | -0,085 | 0,312 | 0,047 |
| **Precuneus** | -0,016 | 0,142 | 0,038 | 0,197 | 0,098 | 0,219 | 0,099 | 0,391 | 0,376 | | 0,148 | 0,203 | 0,141 | 0,102 | 0,032 |
| **Planum temporale** | 0,166 | 0,138 | 0,037 | -0,099 | -0,342 | 0,319 | 0,260 | -0,156 | 0,036 | 0,148 | | 0,003 | 0,328 | 0,265 | 0,168 |
| **Superior frontal gyrus** | 0,023 | -0,197 | 0,331 | 0,039 | 0,048 | 0,037 | 0,383 | 0,125 | 0,030 | 0,203 | 0,003 | | 0,083 | 0,125 | 0,197 |
| **Superior temporal gyrus** | 0,165 | 0,211 | 0,452 | -0,302 | -0,157 | 0,165 | 0,346 | 0,056 | -0,085 | 0,141 | 0,328 | 0,083 | | -0,105 | 0,464 |
| **Temporal pole** | 0,498 | 0,136 | 0,079 | 0,211 | 0,149 | 0,443 | 0,269 | -0,124 | 0,312 | 0,102 | 0,265 | 0,125 | -0,105 | | 0,171 |
| **Triangular part inferior frontal gyrus** | 0,209 | 0,093 | 0,177 | -0,161 | 0,016 | 0,104 | 0,077 | -0,107 | 0,047 | 0,032 | 0,168 | 0,197 | 0,464 | 0,171 | |

**Supplementary table 4C. Pearson correlation coefficients in PD-sym of brain regions identified in cluster 1**

| | Amygdala | Cerebellum white matter | Hippocampus | Putamen | Anterior orbital gyrus | Central operculum | Entorhinal area | Inferior temporal gyrus | Lingual gyrus | Precuneus | Planum temporale | Superior frontal gyrus | Superior temporal gyrus | Temporal pole | Triangular part inferior frontal gyrus |
|---|---|---|---|---|---|---|---|---|---|---|---|---|---|---|---|
| **Amygdala** | | 0,286 | 0,258 | 0,063 | 0,287 | 0,256 | 0,586 | -0,126 | 0,065 | 0,008 | -0,105 | -0,024 | 0,020 | 0,378 | 0,022 |
| **Cerebellum white matter** | 0,286 | | 0,164 | 0,102 | 0,032 | -0,016 | 0,139 | -0,026 | 0,126 | -0,135 | 0,419 | -0,155 | -0,130 | 0,028 | -0,030 |
| **Hippocampus** | 0,258 | 0,164 | | 0,243 | 0,129 | 0,058 | 0,427 | -0,128 | -0,117 | -0,039 | 0,064 | 0,158 | -0,014 | 0,115 | 0,143 |
| **Putamen** | 0,063 | 0,102 | 0,243 | | 0,180 | -0,050 | -0,028 | 0,244 | -0,092 | -0,087 | 0,068 | -0,223 | 0,258 | 0,081 | -0,107 |
| **Anterior orbital gyrus** | 0,287 | 0,032 | 0,129 | 0,180 | | -0,068 | 0,171 | 0,326 | 0,036 | 0,201 | -0,130 | -0,054 | 0,277 | 0,246 | 0,211 |
| **Central operculum** | 0,256 | -0,016 | 0,058 | -0,050 | -0,068 | | 0,384 | -0,372 | 0,318 | -0,043 | -0,229 | -0,033 | 0,047 | 0,385 | -0,095 |
| **Entorhinal area** | 0,586 | 0,139 | 0,427 | -0,028 | 0,171 | 0,384 | | -0,265 | 0,000 | 0,105 | -0,110 | -0,052 | 0,048 | 0,316 | 0,116 |
| **Inferior temporal gyrus** | -0,126 | -0,026 | -0,128 | 0,244 | 0,326 | -0,372 | -0,265 | | -0,032 | -0,068 | 0,003 | -0,193 | 0,052 | 0,018 | 0,077 |
| **Lingual gyrus** | 0,065 | 0,126 | -0,117 | -0,092 | 0,036 | 0,318 | 0,000 | -0,032 | | -0,081 | 0,129 | -0,084 | 0,003 | 0,138 | -0,215 |
| **Precuneus** | 0,008 | -0,135 | -0,039 | -0,087 | 0,201 | -0,043 | 0,105 | -0,068 | -0,081 | | 0,180 | -0,342 | 0,203 | -0,064 | 0,346 |
| **Planum temporale** | -0,105 | 0,419 | 0,064 | 0,068 | -0,130 | -0,229 | -0,110 | 0,003 | 0,129 | 0,180 | | -0,151 | 0,161 | -0,122 | 0,000 |
| **Superior frontal gyrus** | -0,024 | -0,155 | 0,158 | -0,223 | -0,054 | -0,033 | -0,052 | -0,193 | -0,084 | -0,342 | -0,151 | | -0,182 | -0,033 | 0,076 |
| **Superior temporal gyrus** | 0,020 | -0,130 | -0,014 | 0,258 | 0,277 | 0,047 | 0,048 | 0,052 | 0,003 | 0,203 | 0,161 | -0,182 | | 0,266 | 0,024 |
| **Temporal pole** | 0,378 | 0,028 | 0,115 | 0,081 | 0,246 | 0,385 | 0,316 | 0,018 | 0,138 | -0,064 | -0,122 | -0,033 | 0,266 | | 0,083 |
| **Triangular part inferior frontal gyrus** | 0,022 | -0,030 | 0,143 | -0,107 | 0,211 | -0,095 | 0,116 | 0,077 | -0,215 | 0,346 | 0,000 | 0,076 | 0,024 | 0,083 | |
